# Natural history of serum creatine kinase levels and motor, pulmonary, and cardiac functions in 337 patients with Duchenne muscular dystrophy: a retrospective study at a single referral center in Japan

**DOI:** 10.1101/2022.10.19.22281246

**Authors:** Hiroyuki Awano, Yoshinori Nambu, Chieko Ito, Akihiro Kida, Tetsushi Yamamoto, Tomoko Lee, Yasuhiro Takeshima, Kandai Nozu, Masafumi Matsuo

**Author notes:** Corresponding author: Hiroyuki Awano, Organization for Research Initiative and Promotion, Tottori University; 86 Nishi-cho, Yonago, Tottori 6838503, Japan.

## Abstract

**Introduction/Aims:** Duchenne muscular dystrophy (DMD) presents with skeletal muscle weakness, followed by respiratory and cardiac muscle involvement. Recently, with the development of treatments, the need for a natural history to serve as a control for determining treatment efficacy in clinical trials has increased dramatically, however, few large-scale studies have investigated changes in these symptoms. The present study examined the natural history of Japanese DMD patients as a whole and individual patient with genetic mutations eligible for exon skipping therapy.

**Methods:** Medical records of 337 patients with DMD who visited Kobe University Hospital over a period of 30 years from their first visit until 20 years of age were examined.

**Results:** Serum creatine kinase levels showed a stair-step pattern of decline, with extremely high values until 6 and a rapid decline from 7 to 12 years of age. Both the median 10-meter run/walk velocity and rise-from-floor velocity peaked at the age of 4 years and declined with age. The values for respiratory function declined from the age of 11 years. The median left ventricular ejection fraction was >60% until the age of 12 years and rapidly declined from 13 to 15 years of age. Examination of the relationship between gene mutations eligible for exon-skipping therapy and natural history revealed no characteristic findings.

**Discussion:** We found that creatine kinase levels and motor, respiratory, and cardiac functions each exhibited unique changes over time. These findings will be useful in developing new therapeutic agents for DMD and in determining their efficacy in clinical trials.

## Introduction

Duchenne muscular dystrophy (DMD) is a progressive form of muscular atrophy with X-linked inheritance and leads to inevitable death at a young age. Following a symptom-free period after birth, patients with DMD begin to exhibit symptoms of skeletal muscle weakness at approximately 4–5 years of age, including slowed walking velocity.^1^ Serum creatine kinase (CK) levels are extremely high even during the asymptomatic period and serve as a strong clue for the diagnosis of DMD.^2^ Symptoms of muscle weakness worsen with age, and children gradually become unable to independently walk by the age of 12 years.^1^ Muscle weakness progresses with age, and respiratory or cardiac muscle involvement additionally develops. The worsening of these complications is the main cause of mortality at a young age.^3^ Until the 1990s, survival beyond the age of 20 years was difficult to achieve; however, the average life expectancy of patients with DMD born after 1990 is now 28.1 years.^4^ This increase in survival is largely attributable to advances in ventilatory therapy for pulmonary involvement. Nonetheless, cardiac dysfunction remains a major cause of death in DMD, with no effective strategy to overcome it.^2^ Globally, DMD affects one in every 5,000 live births among boys.^5^ Establishing a treatment for DMD is a challenge shared by all humankind. The fundamental therapy for DMD is to re-express defective dystrophin, and several studies have been conducted toward this end. In particular, treatment with antisense oligonucleotide (ASO) to induce exon skipping, which we proposed and reported the first of a treated case, has attracted considerable attention as the most promising therapeutic treatment for DMD.^6^ Consequently, ASO is being aggressively developed, and more than five ASO agents are currently being administered as either approved or investigational drugs for patients with DMD.

The efficacy of therapeutic agents for DMD has also been evaluated by double-blind studies against placebo.^7^ Considering the extremely small number of patients with target gene mutations in double-blind studies using exon-skipping agents, the natural history of patients with DMD may serve as a control to evaluate the efficacy of therapeutic agents.^8^ Hence, the need for a natural history of DMD has been greatly emphasized, and natural history data are being accumulated.^9, 10^ However, to the best of our knowledge, no large-scale studies from Japan have investigated the natural history of DMD. Since the *DMD* gene was cloned as the responsible gene for DMD, we have been actively conducting genetic diagnosis of patients with DMD. Over a period of approximately 30 years, we have identified genetic abnormalities in more than 400 patients with DMD at a single institution in Japan. Subsequently, we proposed and implemented exon-skipping therapy with ASO, paving the way for the establishment of a treatment for DMD.^6, 11, 12^ Furthermore, by utilizing the medical data of several patients, we identified genetic variants in the *DMD* gene that cause a short stature^13^ and late-onset cardiac dysfunction,^14^ as well as a genotype in the *ACTN3* gene that modifies the onset of cardiomyopathy in patients with DMD.^15^ Additionally, we revealed that the motor performance of patients with DMD is related to lower-limb mobility.^16^ In this study, we examined the medical records of each patient with DMD from the first visit until 20 years of age, which were the source of the above-described research results. Subsequently, we extracted the serum CK levels, 10-meter run/walk test results and rise-from-floor test results for motor function evaluation, % forced vital capacity (%FVC) and % forced expiratory volume in 1 second (%FEV1) values for respiratory function evaluation, and left ventricular ejection fraction (LVEF) and left ventricular end-diastolic diameter (LVDd) values for cardiac function evaluation; their changes over time were then examined. Additionally, the natural history of patients with *DMD* gene mutations eligible for exon-skipping therapy for DMD was examined individually. The obtained findings will be useful in developing and designing new therapeutic agents for DMD and in determining their efficacy in clinical trials.

## Materials and Methods

### Study design and ethical considerations

The study design was retrospective and cohort-based. The present study was approved by the Ethics Committee of Kobe University Graduate School of Medicine (approval no.: 1534). This study was conducted in accordance with the principles embodied in the Declaration of Helsinki and the Ethical Guidelines for Human Genome and Genetic Analysis Research. Informed consent was obtained from either the patients or their parents.

### Patients

From March 1991 to March 2019, a total of 439 patients were registered as having DMD in the medical records of the Department of Pediatrics at Kobe University Hospital, located in western Japan, over a period of approximately 30 years. The diagnosis of DMD was confirmed by identifying mutations in the *DMD* gene.^17^ Five patients whose *DMD* gene mutations had major deletions resulting in chromosomal aberrations or adjacent gene aberration syndromes were excluded from this study. Out of the remaining 434 patients, 337 with at least one data point between 0 and 20 years of age for the current evaluation were included in the present study. For patients who participated in a clinical trial involving interventions such as exon skipping, the data obtained after participating in that trial were not used.

Because blood tests, pulmonary function tests, and cardiac function tests are performed once a year in patients with DMD up to 12 years of age and twice a year in patients older than 12 years, the data were collected at least 6 months apart. Exercise function tests are usually performed every 3–6 months; hence, the data were collected at least 3 months apart.

### Serum CK levels

Serum CK levels were measured by Accurus Auto CK (Shino-Test Corporation, Tokyo, Japan) until February 22, 2004, and Cygnus Auto CK (Shino-Test Corporation, Tokyo, Japan) thereafter using a BioMajesty™ analyzer (JCA-BM8040; JEOL Ltd., Tokyo, Japan). The measured values of Accurus Auto CK were converted using the following formula according to the manufacturer’s data: 1.005 × measured value of Accurus Auto CK + 3.2.

### Motor function evaluation

For the 10-meter run/walk test, the patients were instructed to run or walk 10 meters as fast as possible from a starting line to a point 1–2 m ahead of a goal line, where a physical therapist was standing. The time required to run/walk 10 meters was measured using a stopwatch.

The rise-from-floor test measured the time taken to stand up after assuming the supine position on the floor according to the North Star Ambulatory Assessment (NSAA) guidelines.

Both tests were conducted once during each testing session, and the 10-meter run/walk velocity and rise-from-floor velocity was determined from the 10-meter run/walk test time and rise-from-floor test time, respectively. The result was set to zero if a test could not be completed.

### Pulmonary function evaluation

Pulmonary function tests were performed using a spirometer (Autospirometer System 21; Minato Medical Science Co., Ltd., Osaka, Japan), excluding children aged <5 years and patients aged >6 years who were unable to cooperate with in the tests. %FVC and %FEV1 were calculated using standard reference sets.^18, 19^ A nurse measured the height (in millimeters) in the standing position using a stadiometer (Yamato Scale Co., Ltd., Akashi, Japan). For patients with postural problems, height was measured to the nearest millimeter in the supine position using a non-stretch flexible tape. Data from the day of respiratory examination were used for height. If height measurements were not available on the same day, measurements taken within 3 months were used.

### Cardiac function evaluation

Echocardiography was performed in the supine position using a commercially available echocardiography system (Aplio XG; Toshiba Medical Systems, Tochigi, Japan). Three consecutive beats of digital grayscale 2D cine loops were routinely acquired from paratracheal long-axis, short-axis, and standard apical images.^14^ LVEF was assessed using the modified Simpson method; cardiac dysfunction was defined as LVEF <53%.^20^

### Patient grouping by *DMD* gene mutations

For the examination of the relationship between *DMD* gene mutations and natural history, patients with DMD were divided into five groups according to genetic abnormalities targeted by exon-skipping therapy. That is, patients with genetic abnormalities that could be treated by exon skipping of exons 44, 45, 51, and 53 were designated as the exon 44, 45, 51, and 53 groups, respectively, whereas other patients were designated as the “other” group. The exon 44, 45, 51, and 53 groups included deletions of the exons 3–43, 34–43, 45, 45–54, and 45–56; deletions of the exons 10–44, 44, 46–47, 46–48, 46–49, 46–51, 46–53, and 46–55; deletions of the exons 45–50, 48–50, 49–50, 50, and 52; and deletions of the exons 21–52, 45–52, 48–52, 49–52, and 52, respectively.

### Statistical analysis

For each of the extracted data, the minimum, lower quartile, median, upper quartile, and maximum values at each age are shown by box plots. Frequencies are expressed as percentages. All analyses were performed using GraphPad Prism 7.04 (GraphPad Software. Boston. USA).

## Results

### Patients

Out of 337 patients with DMD for whom data were obtained from medical records, 187 (55.5%), 35 (10.4%), and 71 (21.1%) were identified as having deletions, duplications, and nonsense mutations as *DMD* gene abnormalities, respectively. Diverse mutations were identified in the remaining patients. Among 187 patients with exon deletions, 17, 34, 32, and 36 patients were in the exon 44, 45, 51, and 53 groups, respectively, who were eligible for treatment by exon skipping of exons 44, 45, 51, and 53; on the other hand, 226 patients were in the “other” group. However, eight patients with a single deletion of exon 52 were in both the exon 51 and 53 groups.

Of the 337 patients, 52 (15.4%) received oral steroid therapy. Steroid dosage and administration varied widely from individual to individual; dosage was 1∼25 mg/day, administration was daily, every other day, or 10 days-on and 20 days-off. A total of 64 (19.0%) patients received medications for cardiomyopathy such as angiotensin-converting enzyme inhibitors, angiotensin receptor blockers, and beta-blockers. Overall, 9 (2.6%) out of the 337 patients died prior to the age of 21 years.

### Serum CK levels

Out of the 337 patients, 331 underwent serum CK level measurement a total of 1792 times. The median CK value was in the upper 10,000 U/L range from 0 to 5 years of age, reached approximately 16,000 U/L at the age of 6 years, sharply dropped to 3,749 U/L at the age of 11 years, subsequently decreased to 2,839 U/L at the age of 12 years, and remained relatively low in the thousands or hundreds thereafter (Figure 1). The CK values showed a staircase-like change over time, indicating an age-dependent disease progression.

**Figure 1.**
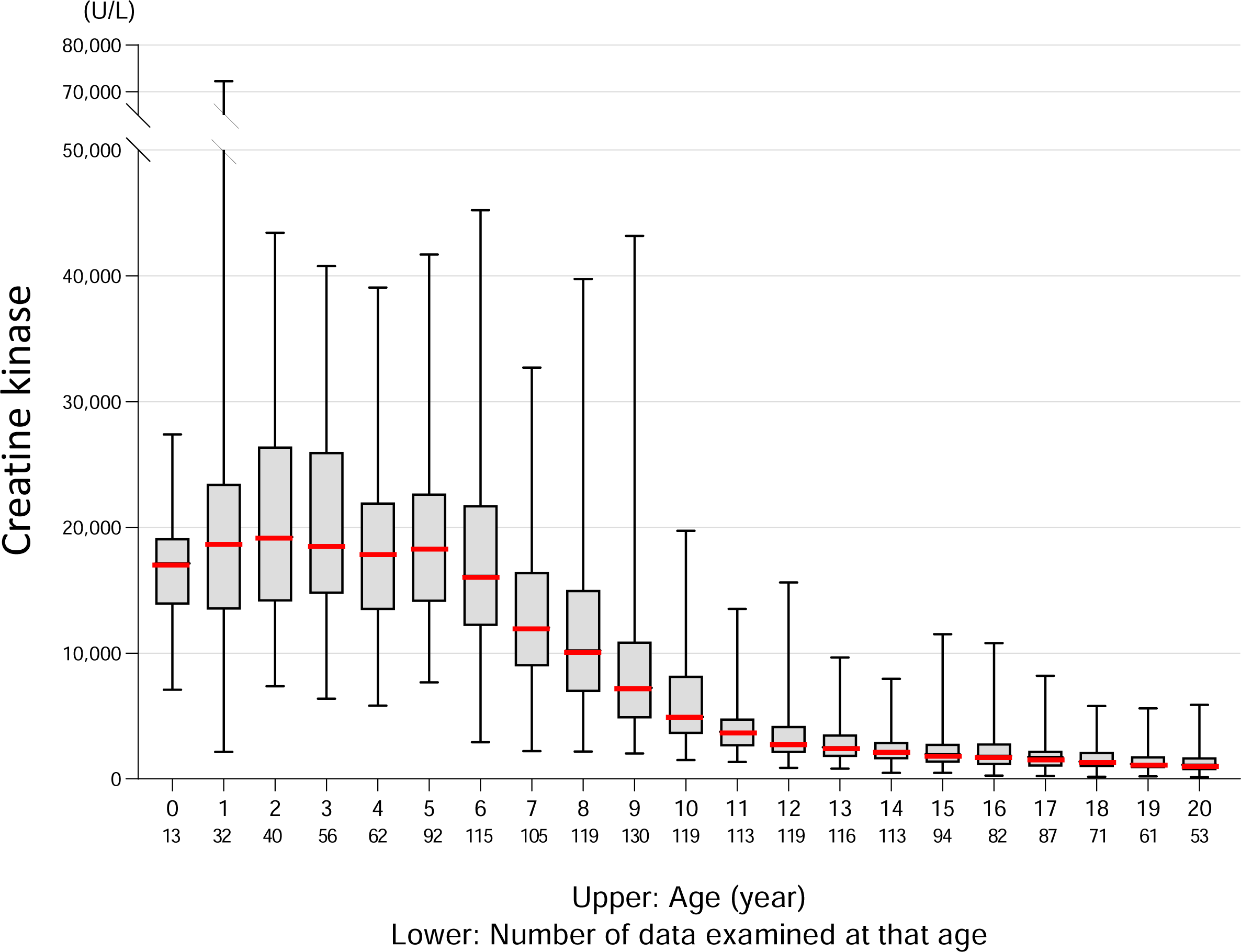
Serum CK levels over time Box-and-whisker plots for serum CK levels in each age group from 0 to 20 years are shown. The median value is marked with a thick red line. The number of CK measurements at each age is noted below. CK, creatine kinase.

### Motor function evaluation

Among the 337 patients with DMD, 118 underwent at least one motor function evaluation, for a total of 1234 and 1218 attempts of the 10-meter run/walk test and rise-from-floor test, respectively. All tested patients completed the 10-meter run/walk test until the age of 5 years; however, failure cases began to appear at the age of 6 years, and the percentage of failure cases then increased with increasing age (Figure 2A). At the age of 10 years, the percentage of failure exceeded 50%.

**Figure 2.**
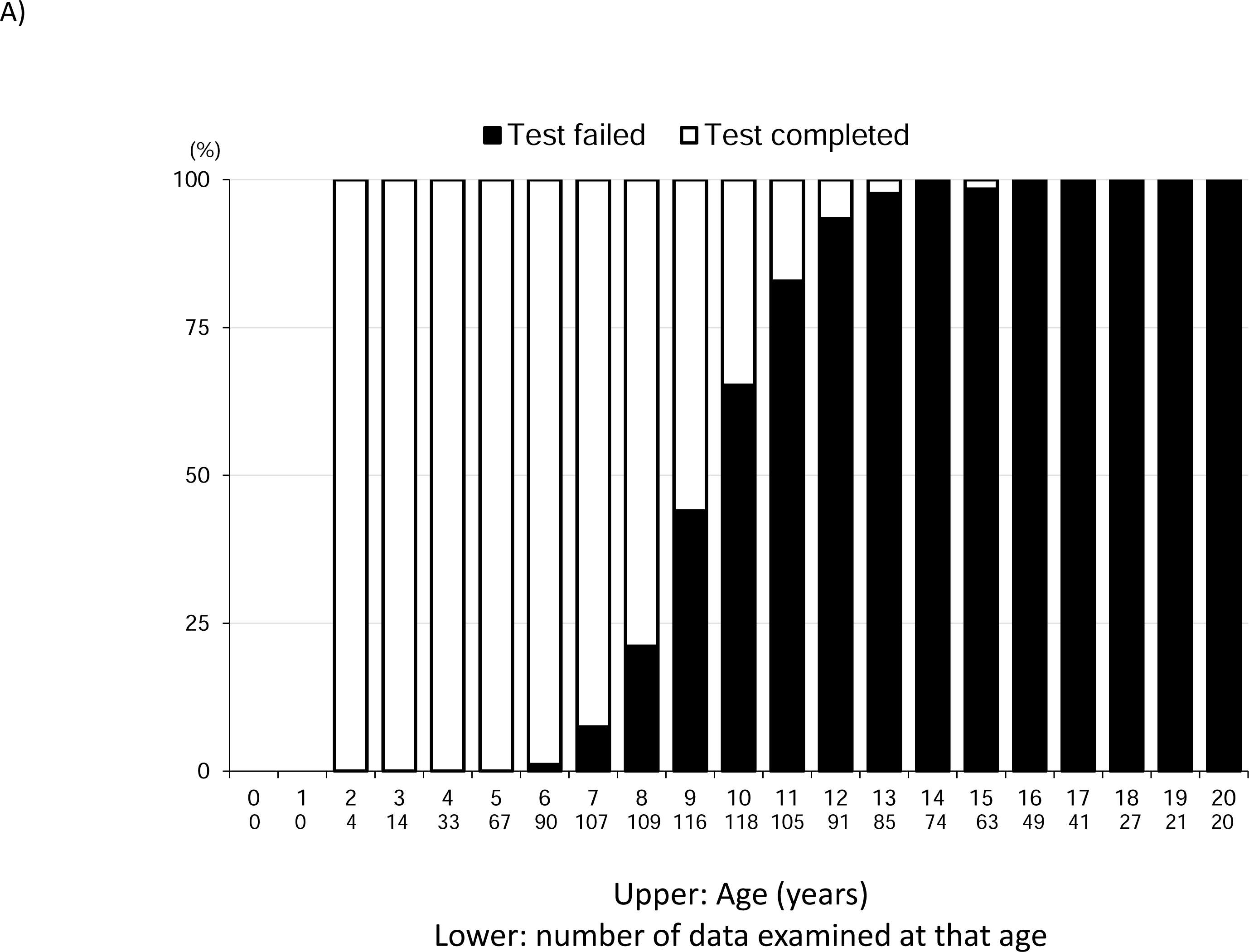
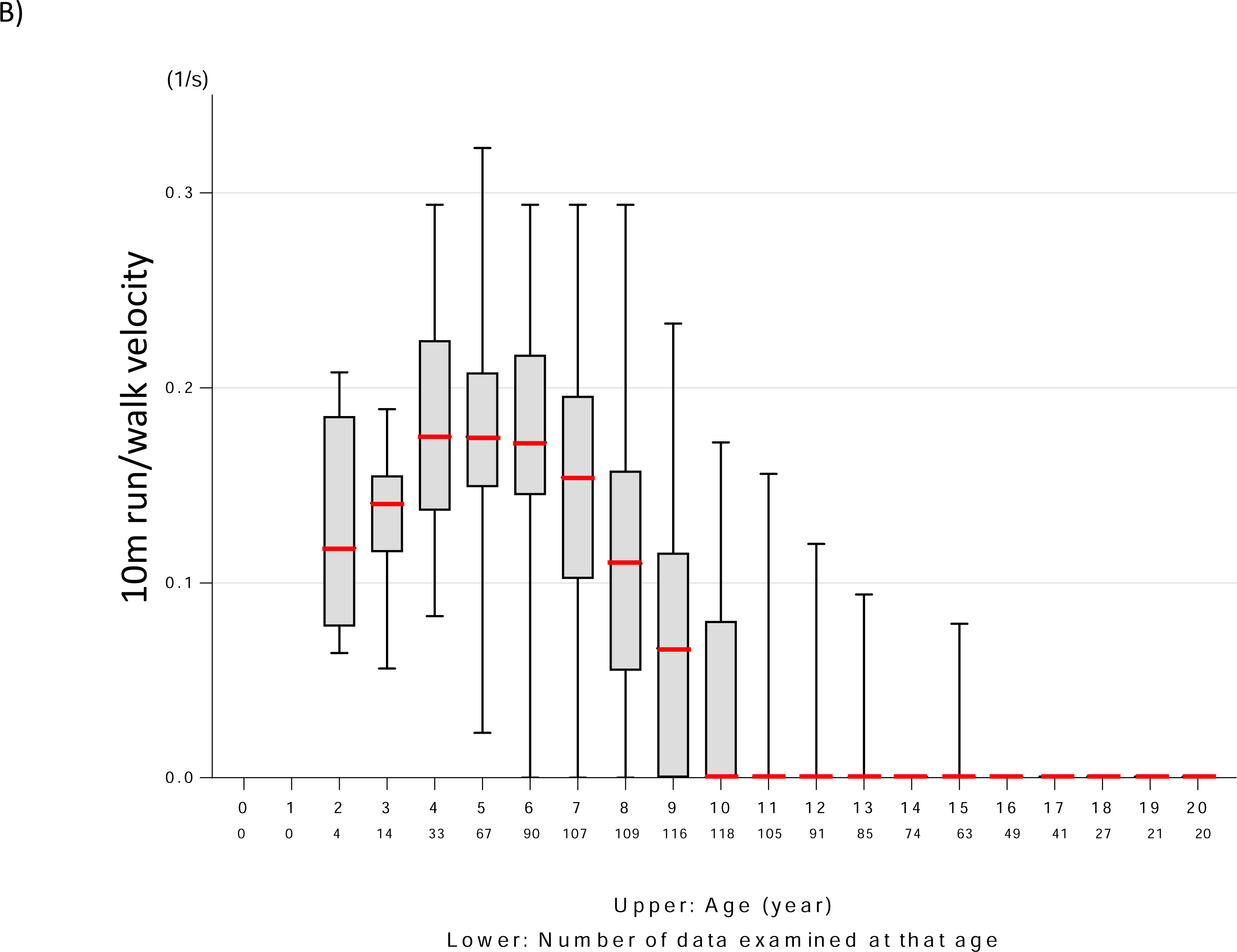
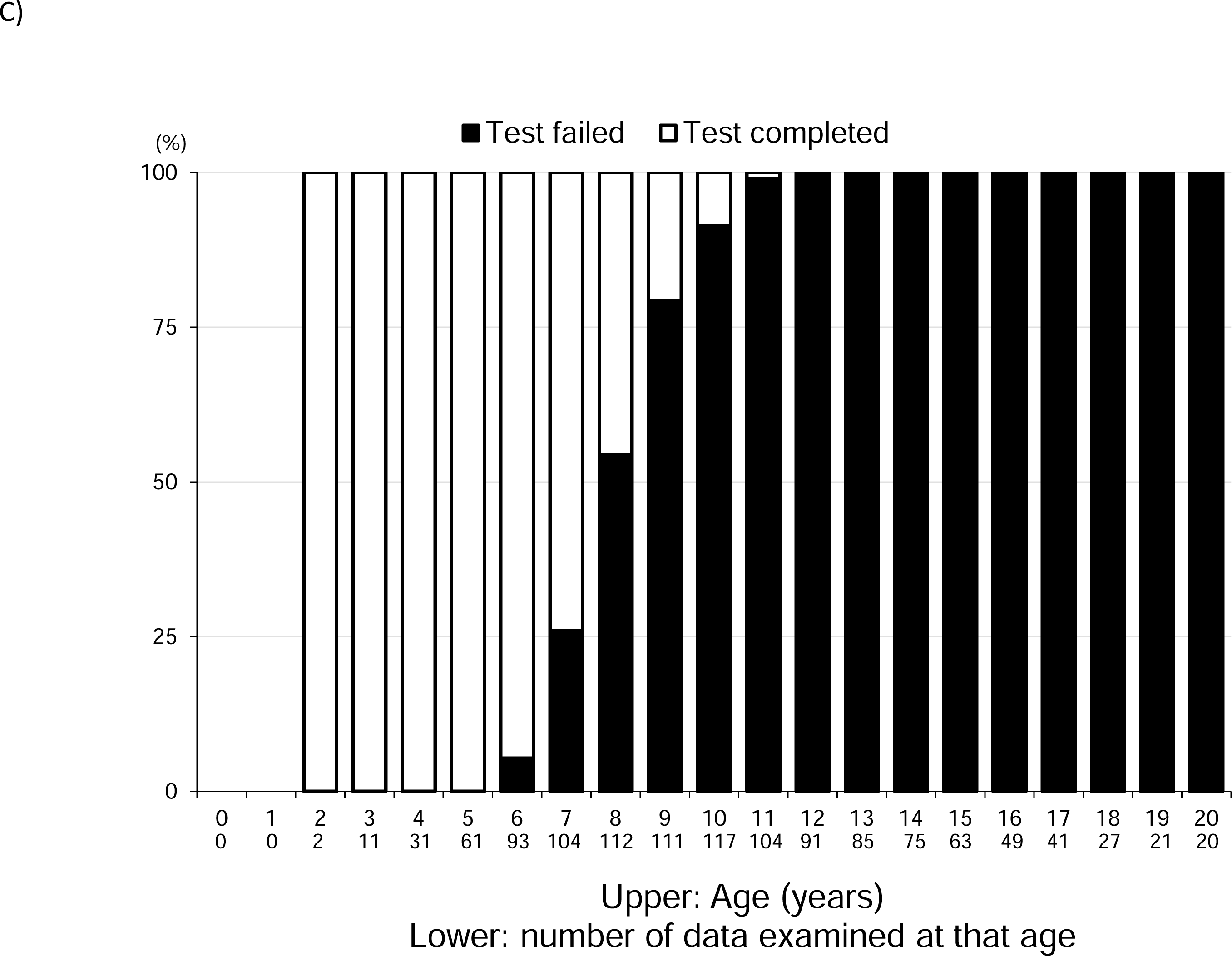
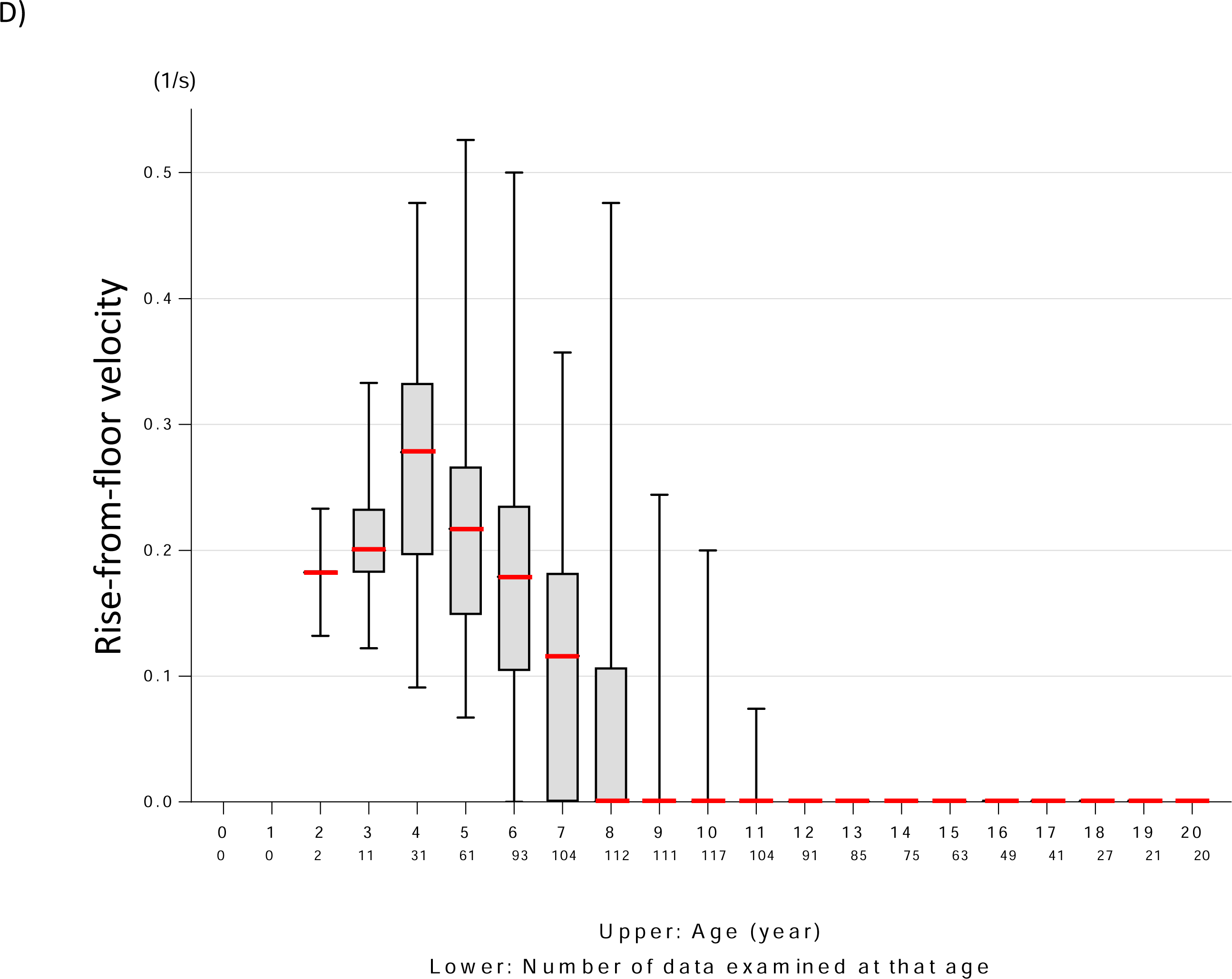
Results of motor function evaluation A. Percentage of failure in the 10-meter run/walk test according to age The percentage of instances of non-achievement in the 10-meter run/walk test is shown in black boxes. B. Changes in 10-meter run/walk velocity over time The median and box whiskers are shown for each age group based on the 10-meter run/walk velocity. The median is marked with a thick red line. C. Percentage of failure to complete the rise-from-floor test according to age The percentage of instances of non-achievement in the rise-from-floor test is shown in black boxes. D. Changes in rise-from-floor velocity over time The median and box whiskers are shown for each age group based on the rise-from-floor velocity. The median is marked with a thick red line.

We subsequently examined the age trend for 10-meter run/walk velocity (1/s) (Figure 2B). The median increased from 2 to 4 years of age, reached the maximum level at 4 years of age, and remained almost constant from 4 to 6 years of age. The median started to decline at the age of 7 years and reached zero at the age of 10 years.

The percentage of patients who failed on the rise-from-floor test was examined by age groups (Figure 2C). All patients were able to rise from the floor until the age of 5 years; however, failure cases began to appear at the age of 6 years. More than half of the patients were unable to rise from the floor at the age of 8 years, and all patients were unable to rise from the floor at the age of 12 years.

We further examined the evolution of the rise-from-floor velocity (1/s) with age (Figure 2D). The median increased from 2 years of age, and reached the maximum level at 4 years of age (0.28 (1/s)). The median decreased with age from the age of 5 years onward, reaching 0 at the age of 8 years.

### Pulmonary function evaluation

A total of 609 %FVC and 608 %FEV1 results were obtained from 139 patients with DMD. The percentage of lung function tests attempted but not completed was summarized by age (Figure 3A).

**Figure 3.**
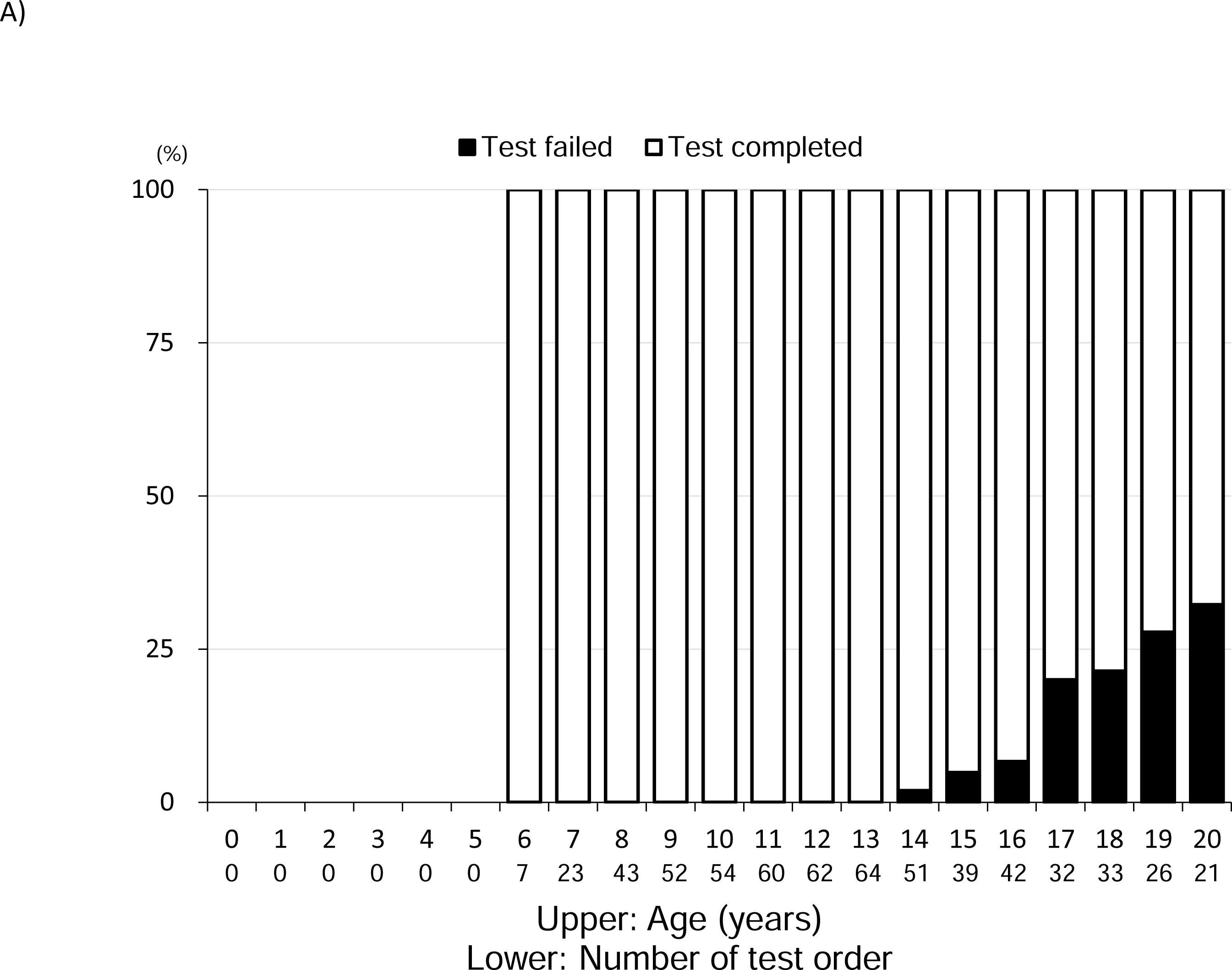
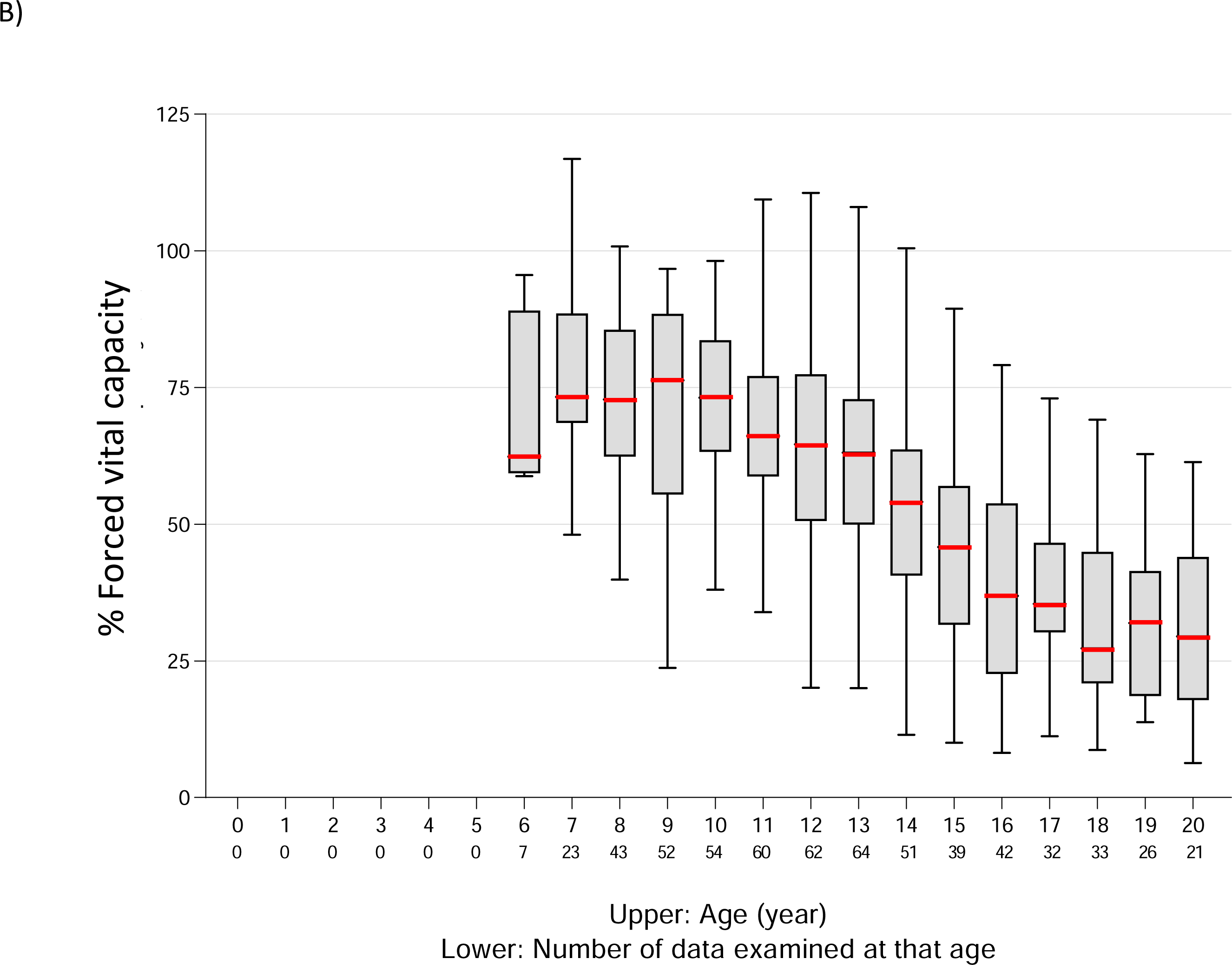
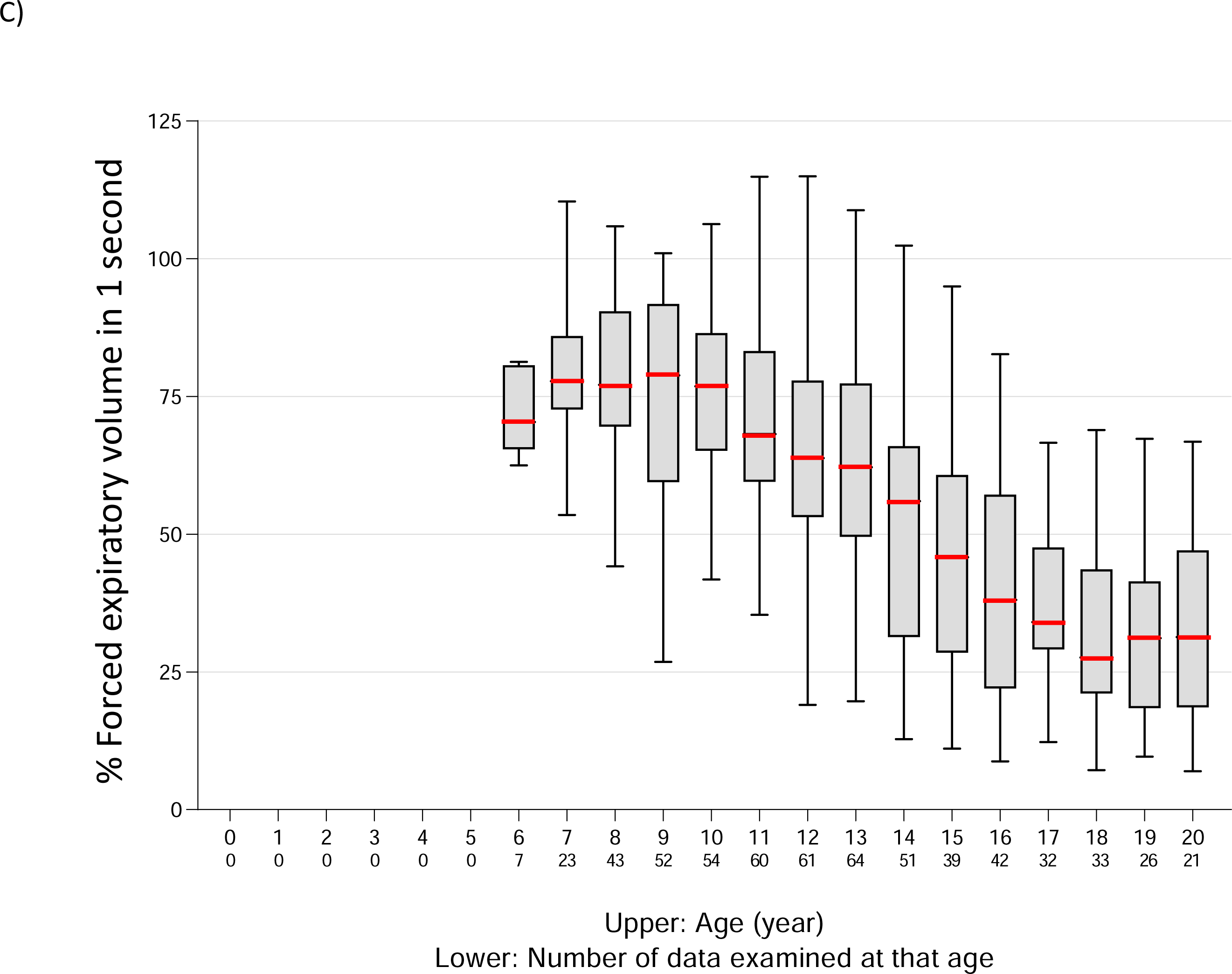
Results of pulmonary function evaluation A. Percentage of respiratory function test failures The black frame indicates failure. B. %FVC values according to age Median and box whiskers are shown for each age group. The median is indicated by a thick red line. C. %FEV1 values according to age Median and box whiskers are shown for each age group. The median is marked with a thick red line. %FEV1, % forced expiratory volume in 1 second; %FVC, % forced vital capacity.

Failed test attempts began to appear at 14 years of age, with the percentage reaching approximately 30% at the age of 20 years. The median %FVC peaked at the age of 9 years and then gradually decreased from the age of 10 to 18 years (Figure 3B). Similar to %FVC, the median %FEV1 peaked at the age of 9 years and gradually decreased from the age of 10 to 18 years (Figure 3C).

### Cardiac function evaluation

Echocardiography was performed in 190 out of the 337 patients, yielding 867 LVEF values and 1016 LVDd values in total. Changes in LVEF values over time were examined (Figure 4A). The median LVEF was above 60% from 3 to 12 years of age but decreased to 52.8% at the age of 13 years and 45.4% at the age of 15 years. Subsequently, it showed a stair-step change from 16 to 20 years of age, with a plateau of approximately 45%.

**Figure 4.**
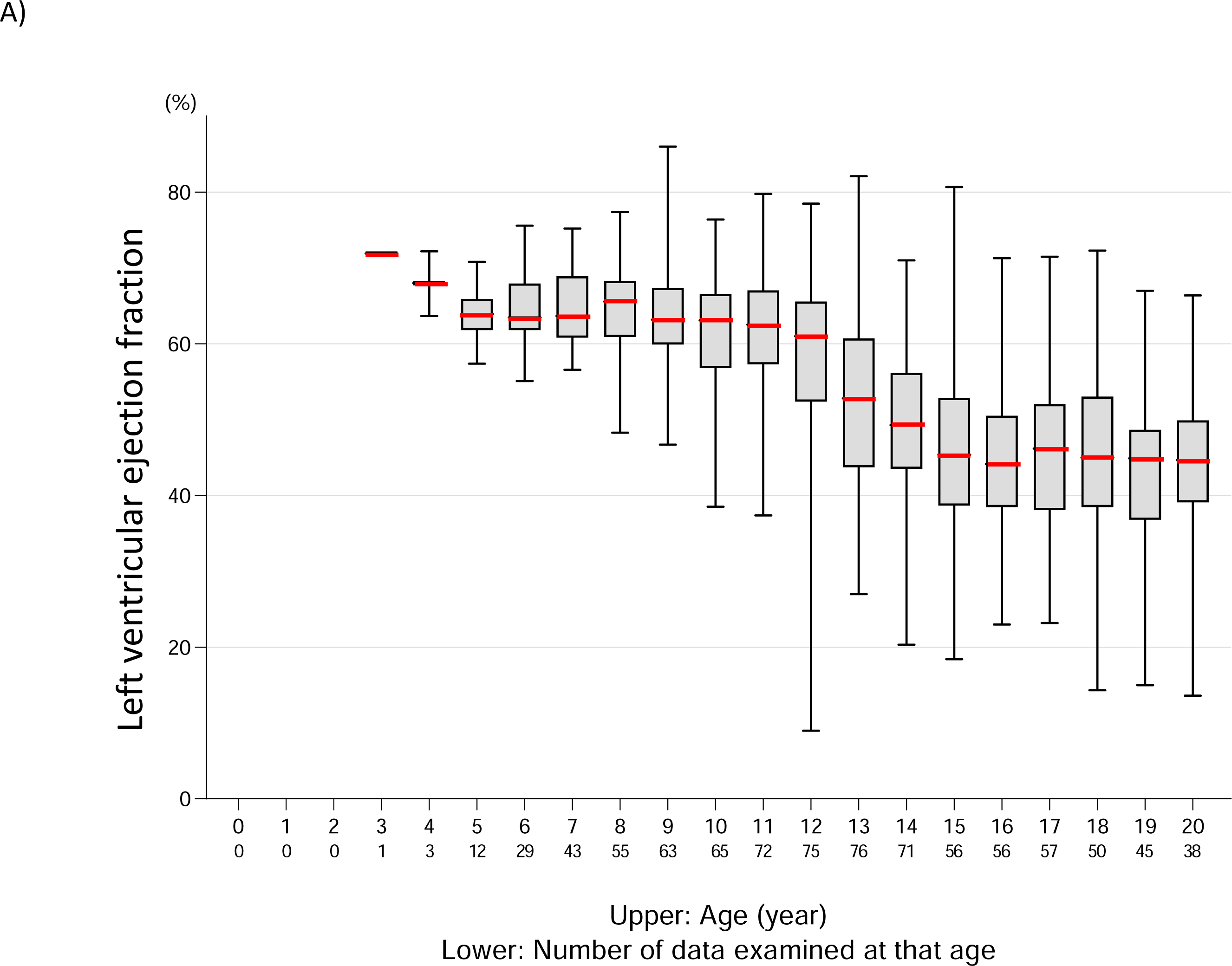
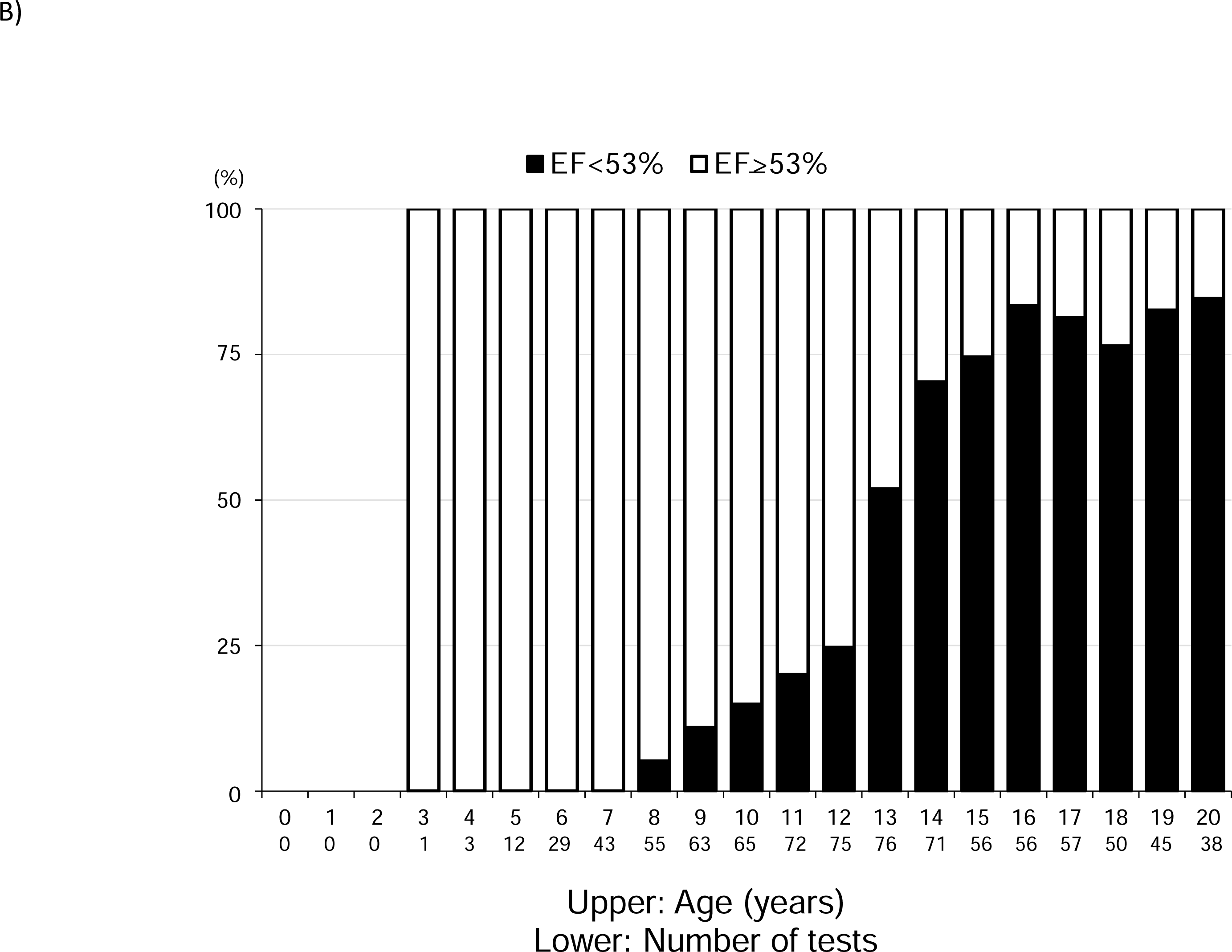
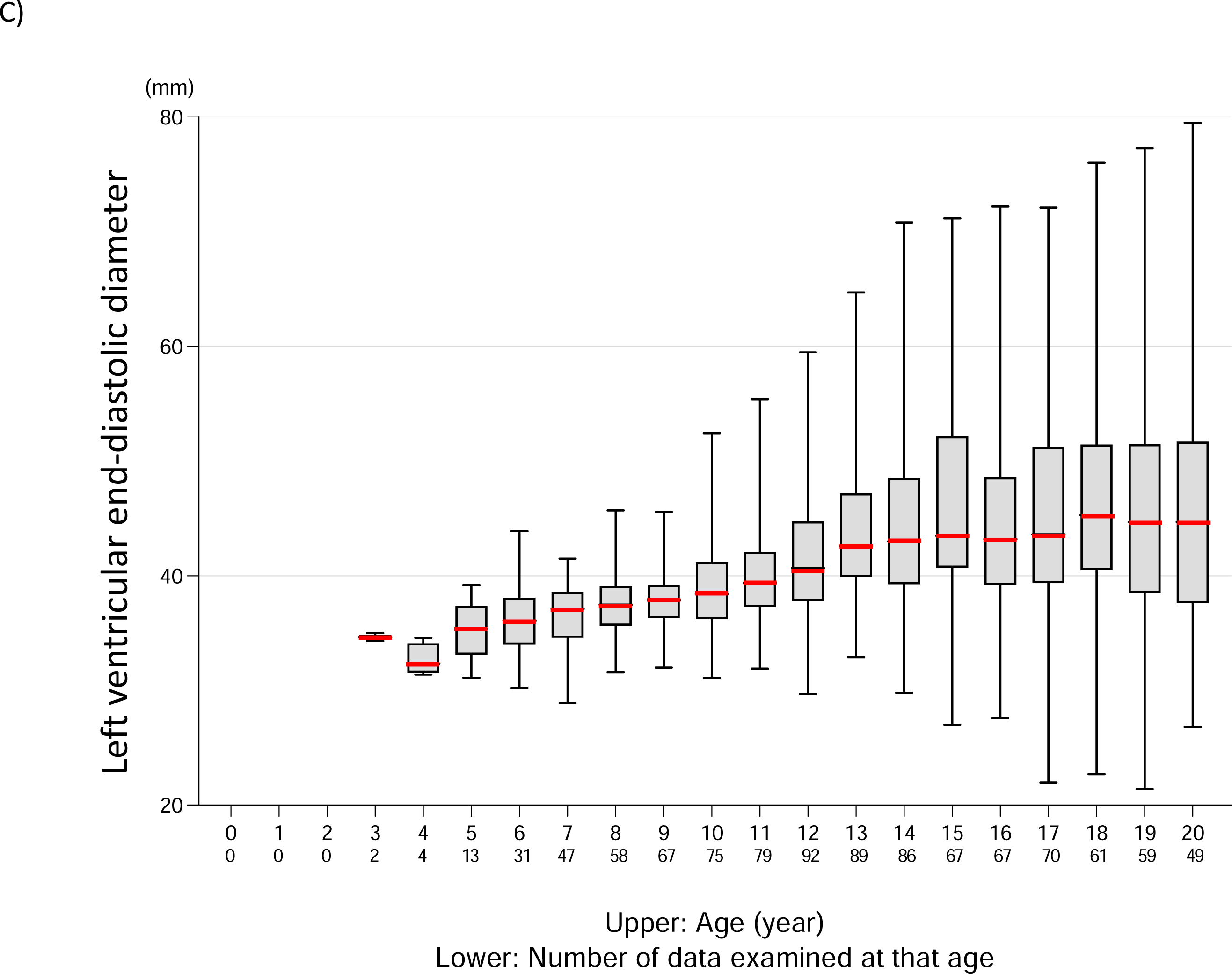
Results of cardiac function evaluation A. LVEF values according to age Median LVEF and box whiskers are shown for each age group. The median values are marked with a thick red line. B. Proportion of cardiac dysfunction according to age The black boxes represent heart failure with LVEF <53%. C. LVDd values according to age Median and box whiskers are shown for each age group. The median is marked with a thick red line. LVDd, left ventricular end-diastolic diameter; LVEF, left ventricular ejection fraction.

We defined cardiac dysfunction as LVEF <53% and examined age-specific rates of cardiac dysfunction (Figure 4B). The first case of heart failure appeared at the age of 8 years. Thereafter, the proportion of those with cardiac dysfunction increased year by year, with more than 50% having cardiac dysfunction at the age of 13 years and more than 75% having cardiac dysfunction at the age of 15 years or later. We also examined the change in LVDd over time (Figure 4C). In each age group, the median value tended to increase with age, starting at 4 years of age.

### *DMD* gene mutation type and natural history

Based on the variant type of *DMD* gene that patients with DMD had, they were classified into four groups according to the number of exons targeted by exon-skipping therapy—namely, the exon 44 group (*n*=17), exon 45 group (*n*=34), exon 51 group (*n*=32), and exon 53 group (*n*=36). The other patients (*n*=226) had other abnormalities. Changes in their data over time were examined.

Examination of patient-specific changes in serum CK levels over time revealed no characteristic pattern of change in each group (Supplementary Figure 1A). In each group, the CK levels decreased to below 10,000 U/L at almost 11 years of age (Supplementary Figure 1B). These results indicated that there existed no clear differences in CK level changes over time according to genetic variant type. Likewise, examination of patient-specific changes in 10-meter run/walk velocity over time showed no major characteristic pattern of change in each group (Supplementary Figure 2A). All patients in the exon 44, 45, 51, and 53 groups failed the 10-meter run/walk test at ages of 12, 12, 13, and 13 years, respectively; on the other hand, all patients in the other group with non-treatment target mutations failed the 10-meter run/walk test at the age of 16 years (Supplementary Figure 2B).

Changes in rise-from-floor velocity over time were also examined in each individual; however, no characteristic findings were observed (Supplementary Figure 2C). The age at which all patients failed the rise-from-floor test was 11 years in all four groups eligible for the exon-skipping therapy but 15 years in the other group (a difference of 4 years) (Supplementary Figure 2D). Functional loss tended to occur later in the other group.

With respect to pulmonary function, no characteristic findings were detected in each group when changes in %FVC and %FEV1 over time were examined in each individual (Supplementary Figure 3A and B). Cases of %FVC <50% were found in all patients in the four groups eligible for the exon-skipping therapy, but not in all patients in the other group, although they appeared from a younger age (Supplementary Figure 3C).

LVEF and LVDd in the cardiac function tests were examined in each group of individuals (Supplementary Figure 4A and B); such examination did not reveal any characteristic findings in each group.

The percentage of patients showing cardiac dysfunction was determined to be 100% at older ages in the exon 45 and 53 groups, but not necessarily 100% in the other group. These results suggested the existence of a relationship between cardiac dysfunction progression and genetic abnormality type.

## Discussion

DMD is characterized by progressive muscle atrophy with early childhood onset. The present study conducted on a Japanese population of patients with DMD showed that the median age at onset of progressive change in each endpoint was different: in particular, serum CK levels started at the age of 6 years, 10 m run/walk test at the age of 7 years, rise-from-floor test at the age of 5 years, %FVC and %FEV1 at the age of 10 years, LVEF at the age of 13 years, and LVDd at the age of 5 years, with a trend toward decline. DMD results from a dystrophin deficiency; nonetheless, the age at which pathological changes caused by this deficiency appear was different among the endpoints, and this should be noted when setting the timing of therapeutic interventions for DMD.

Serum CK is a powerful biomarker for the diagnosis of DMD. The largest age-specific CK analysis, which was conducted on 456 patients with DMD aged 0–23 years, reported that CK levels were the highest at 4–5 years of age and slowly declined between 18–23 years of age.^21^ In the present study, 1792 CK values were obtained from 331 patients, which allowed for a more detailed age-specific CK analysis. Our analysis revealed that the median age-specific CK value declined in a staircase-like fashion, with a steep drop between the ages of 6 and 12 years. This steep decline from 6 to 12 years of age was thought to reflect the reduction in muscle mass during this period. In a recent clinical trial of micro-dystrophin gene therapy for DMD, a decrease in serum CK levels was employed as the most promising biomarker for determining therapeutic efficacy.^22^ On the other hand, our results indicated that serum CK levels showed a stair-step pattern of decline, suggesting that the determination of therapeutic efficacy using CK levels requires a comparison with naturally occurring CK levels that takes age at the time of determination into account. Furthermore, we were able to develop a curve showing the decrease in CK levels for each individual patient, which may be used as a basis for the future evaluation of DMD treatment. We expect that our CK data will be used to determine the efficacy of DMD treatment.

In addition to their ease, the 10-meter run/walk test and rise-from-floor test have utility in the assessment of motor performance in DMD^23^ and have been employed in clinical trials of exon-skipping therapy to evaluate its efficacy.^8^ Therefore, it is expected that the natural history of the 10-meter run/walk test and rise-from-floor test will be more widely used in clinical practice as a method for motor function evaluation in DMD in the future. To the best of our knowledge, comparisons between the type of *DMD* gene abnormalities and the results of the 10-meter run/walk test and rise-from-floor test have not been conducted very often. However, the percentage of patients who could perform the rise-from-floor test has been reported to be greater in the exon 51 and 53 groups than in the exon 44 and 45 groups.^24^ Nevertheless, no clear difference between the groups of genetic abnormalities was found in our study.

Preserving or improving the pulmonary function is one measure in the evaluation of therapeutic agents for DMD. The values of %FVC and %FEV1, which were the two pulmonary function markers in the present study, tended to slowly decrease with age. This trend was consistent with the findings of previous reports.^25, 26^

Cardiac dysfunction is currently the leading cause of mortality in DMD. Attempts to detect myocardial abnormalities in DMD have been made using electrocardiography; however, a wide variety of electrocardiogram abnormalities have been detected. Linking these abnormalities to the pathogenesis of DMD has been difficult.^27, 28^ Biannual echocardiography starting at the age of 10 years is recommended to detect cardiac dysfunction early.^29^ In the present study, a sharp decline in LVEF was observed between the ages of 13 and 15 years, supporting the validity of performing echocardiography starting at the age of 10 years. With respect to the relationship between genotype and the occurrence of cardiac dysfunction, in the present study, 100% of patients in the exon 45 and 53 groups had cardiac dysfunction due to their advanced age; in contrast, this was not always the case in the other group. A relationship between cardiac dysfunction and genetic abnormalities is suggested, and further studies are therefore needed.

Because DMD is primarily characterized by skeletal muscle weakness, the focus of determining the effectiveness of DMD treatment is on motor function improvement. As cardiac and pulmonary complications are the actual causes of death in DMD, attention has recently been directed toward overcoming cardiac and pulmonary complications. It is greatly expected that the results of this natural history elucidation will be utilized in the development of future therapeutic agents.

### Limitations

As the present study is a retrospective study based on medical records, it has the common shortcomings of retrospective analysis methods, such as a lack of uniformity in data. However, given that the data were from a single institution, data variability could be expected to be small. Additionally, the NSAA, which is currently commonly used for motor function assessment in DMD, was not used. In Japanese hospitals, the health insurance system restricts the content of medical care, and conducting a large number of tests during general medical care is difficult. However, the 10-meter run/walk test and rise-from-floor test are relatively easy to administer and have some similarities with the NSAA. In the present cohort, the number of patients taking oral steroids was relatively small because treatment with steroids was largely based on parental preference and several parents did not want their children to take steroids owing to concerns about their side effects. Therefore, the level of statistically significant treatment between steroid-taking and non-taking cases was not reached. Rather, the present results are considered to be the natural history of steroid non-taking cases.

### Conclusions

We analyzed the natural history of patients with DMD in Japan during childhood and adolescence and showed that CK levels and motor, respiratory, and cardiac functions each exhibited unique changes over time. These findings will be useful in developing and designing new therapeutic agents for DMD and in determining their efficacy in clinical trials.

## Data Availability

The data used in this article can be obtained upon reasonable individual request to the corresponding author for their release.

## Acknowledgements

The authors thank Kosuke Kanemoto and Takuya Maoka (KNC Laboratories, Inc., Kobe, Japan) as well as Yuichiro Niwata (Daiichi Sankyo Co., Ltd., Tokyo, Japan) for their help in organizing the data.

## Funding Statement

This work was supported in part by an intramural research grant (29-4) from the Department of Neurological and Psychiatric Disorders, National Center of Neurology and Psychiatry, and by a Grant-in-Aid for Scientific Research (19K20673) and Health and Labor Sciences Research Grant (21FC1006)

## Conflicts of Interest Disclosure

Masafumi Matsuo is a KNC-funded endowed chair at Kobe Gakuin University as well as a medical advisor to Daiichi Sankyo Co., Ltd. (Tokyo, Japan) and JCR Pharmaceuticals Co., Ltd. (Ashiya, Japan). Kandai Nozu received a grant from Zenyaku Kogyo Co., Ltd. (Tokyo, Japan), consulting fees from Toa Eiyo Ltd. (Tokyo, Japan), Kyowa Kirin Co., Ltd. (Tokyo, Japan), and Taisho Pharmaceutical Co. Ltd. (Tokyo, Japan) and lecture fees from Sumitomo Pharma Co., Ltd. (Osaka, Japan), Chugai Pharmaceutical Co. Ltd. (Tokyo, Japan), and Kyowa Kirin Co., Ltd., and has patents for developing exon-skipping therapy for Alport syndrome patients from Daiichi Sankyo Co., Ltd.. Yasuhiro Takeshima received consulting fees from Daiichi Sankyo Co., Ltd. and Nippon Shinyaku Co., Ltd. (Tokyo, Japan), lecture fees from Nippon Shinyaku Co., Ltd., has patent from Daiichi Sankyo Co., Ltd. and participate on Advisory Board. Hiroyuki Awano received consulting fees from Daiichi Sankyo Co., Ltd., Nippon Shinyaku Co., Ltd., and Takeda Pharmaceutical Company Limited (Tokyo, Japan), and lecture fees from Nippon Shinyaku Co., Ltd., Chugai Pharmaceutical Co. Ltd., Pfizer Japan Inc. (Tokyo, Japan) and JCR Pharmaceuticals Co., Ltd. Tomoko Lee received lecture fees from Nippon Shinyaku Co., Ltd.

The remaining authors have no conflicts of interest.

## Ethics Approval Statement

This study was approved by the Ethics Committee of Kobe University Graduate School of Medicine (approval no.: 1534). This study was conducted in accordance with the principles embodied in the Declaration of Helsinki and the Ethical Guidelines for Human Genome and Genetic Analysis Research.

## Patient Consent Statement

Informed consent was obtained from all patients or their parents.

## Ethical Publication Statement

We confirm that we have read the Journal’s position on issues involved in ethical publication and affirm that this report is consistent with those guidelines.

## Abbreviations

%FEV1: % forced expiratory volume in 1 second
%FVC: % forced vital capacity
ASO: antisense oligonucleotide
CK: creatine kinase
DMD: Duchenne muscular dystrophy
NSAA: North Star Ambulatory Assessment
LVDd: left ventricular end-diastolic diameter
LVEF: left ventricular ejection fraction.

## Supplementary figures

Supplementary Figure 1 *DMD* genotype and serum CK levels

A. Individual secular changes in serum CK levels by gene abnormality group

CK values for each individual patient by genetic abnormality group are plotted by age and connected by lines. Individuals are indicated by the color of the lines.

B. Percentage of serum CK levels <10,000 U/L

The percentage of serum CK levels <10,000 U/L at each age is shown for each group.

CK, creatine kinase.

Supplementary Figure 2 *DMD* genotype and motor function evaluation

A. Group-specific changes in 10-meter run/walk velocity over time by genetic abnormality group The 10-meter run/walk velocity in each individual patient by genetic abnormality group is plotted for each age group and connected by lines. Individuals are indicated by the color of the lines.

B. Percentage of noncompliance with the 10-meter run/walk test

The percentage of noncompliance with the test determined for each age group is shown by group.

The black boxes represent the percentage of noncompliance with the test.

C. Changes in rise-from-floor velocity over time by genetic abnormality group

The rise-from-floor velocity in each individual patient by genetic abnormality group is plotted by age and connected by lines. Individuals are indicated by the color of the lines.

D. Percentage of noncompliance with the rise-from-floor test

The percentage of noncompliance with the test determined for each age group is shown by group. The black boxes represent the percentage of noncompliance with the test.

Supplementary Figure 3 *DMD* genotype and pulmonary function evaluation

A. Changes in %FVC over time by genetic abnormality group

The %FVC of patients by genetic abnormality group is plotted by age and connected by lines. Individuals are indicated by the color of the lines.

B. Changes in %FEV1 over time by genetic abnormality group

The %FEV1 of patients by genetic abnormality group is plotted by age and connected by lines. Individuals are indicated by the color of the lines.

C. Percentage of %FVC <50% by genetic abnormality group

The percentage of FVC <50% determined for each age group is shown by group. The black boxes represent the percentage of %FVC <50%.

%FEV1, % forced expiratory volume in 1 second; %FVC, % forced vital capacity.

Supplementary Figure 4 *DMD* genotype and cardiac function evaluation

A. LVEF by genetic abnormality group, by group, and by individual over time

The LVEF of patients by genetic abnormality group is plotted by age and connected by lines. Individuals are indicated by the color of the lines.

B. LVDd by genetic abnormality group, by group, and by individual over time

The LVDd of patients by genetic abnormality group is plotted by age and connected by lines. Individuals are indicated by the color of the lines.

C. Proportion of cardiac dysfunction by genetic abnormality group The black boxes represent the percentage of cardiac dysfunction.

LVDd, left ventricular end-diastolic diameter; LVEF, left ventricular ejection fraction.

## Notes

### Summary of Updates

Abstruct and main text revised to conform to styles of the peer-review journal to which we plan to submit this manuscript; Supplementary figures revised; Author affiliations updated; Corresponding author information updated.

