## Supplementary Figures for "Natural history of serum creatine kinase levels and motor, pulmonary, and cardiac functions in 337 patients with Duchenne muscular dystrophy: a retrospective study at a single referral center in Japan"

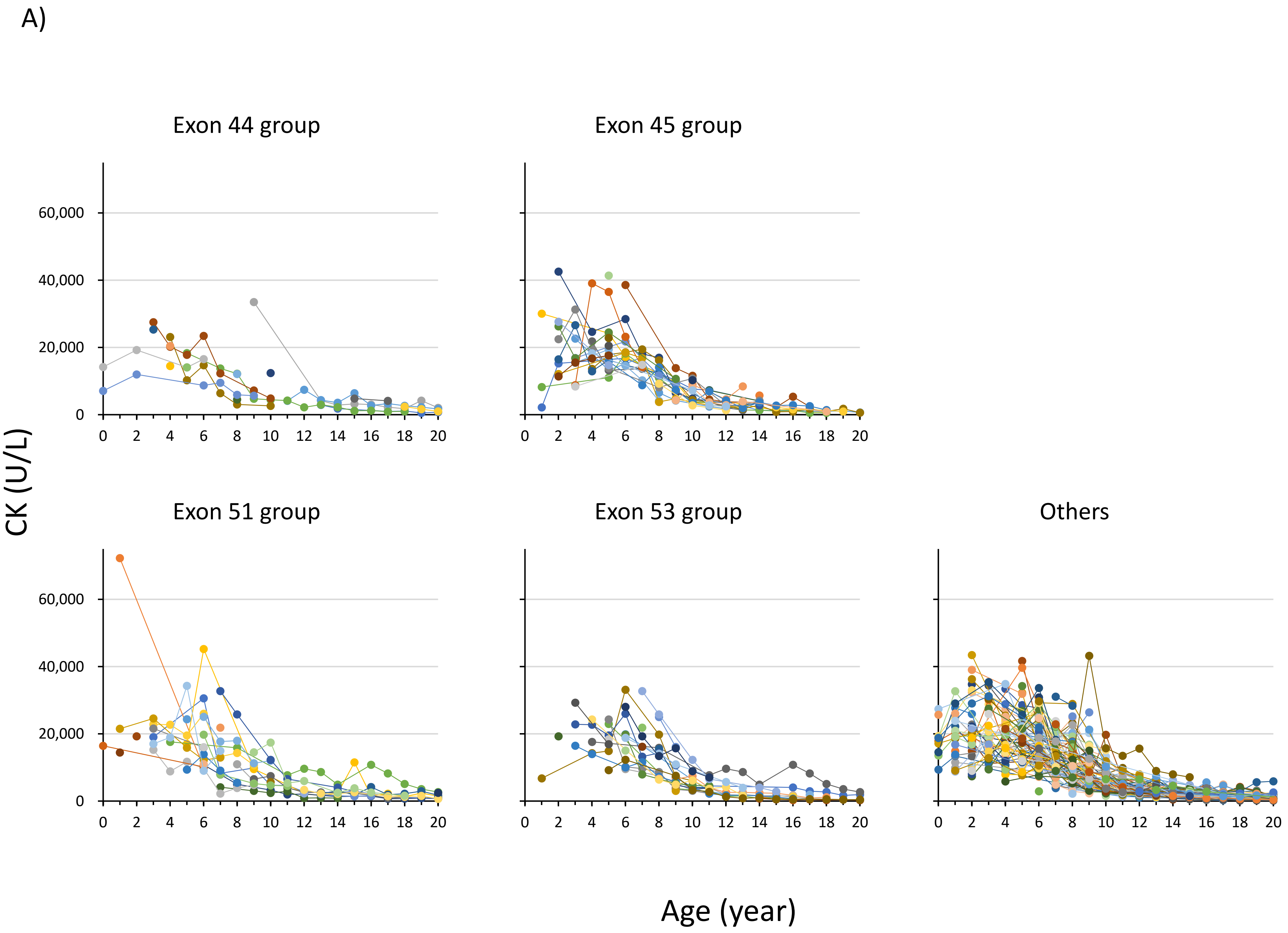
A)
Exon 44 group
Exon 45 group
Exon 51 group
Exon 53 group
Others
CK (U/L)
Age (year)
60,000
40,000
20,000
0
0 2 4 6 8 10 12 14 16 18 20

B)

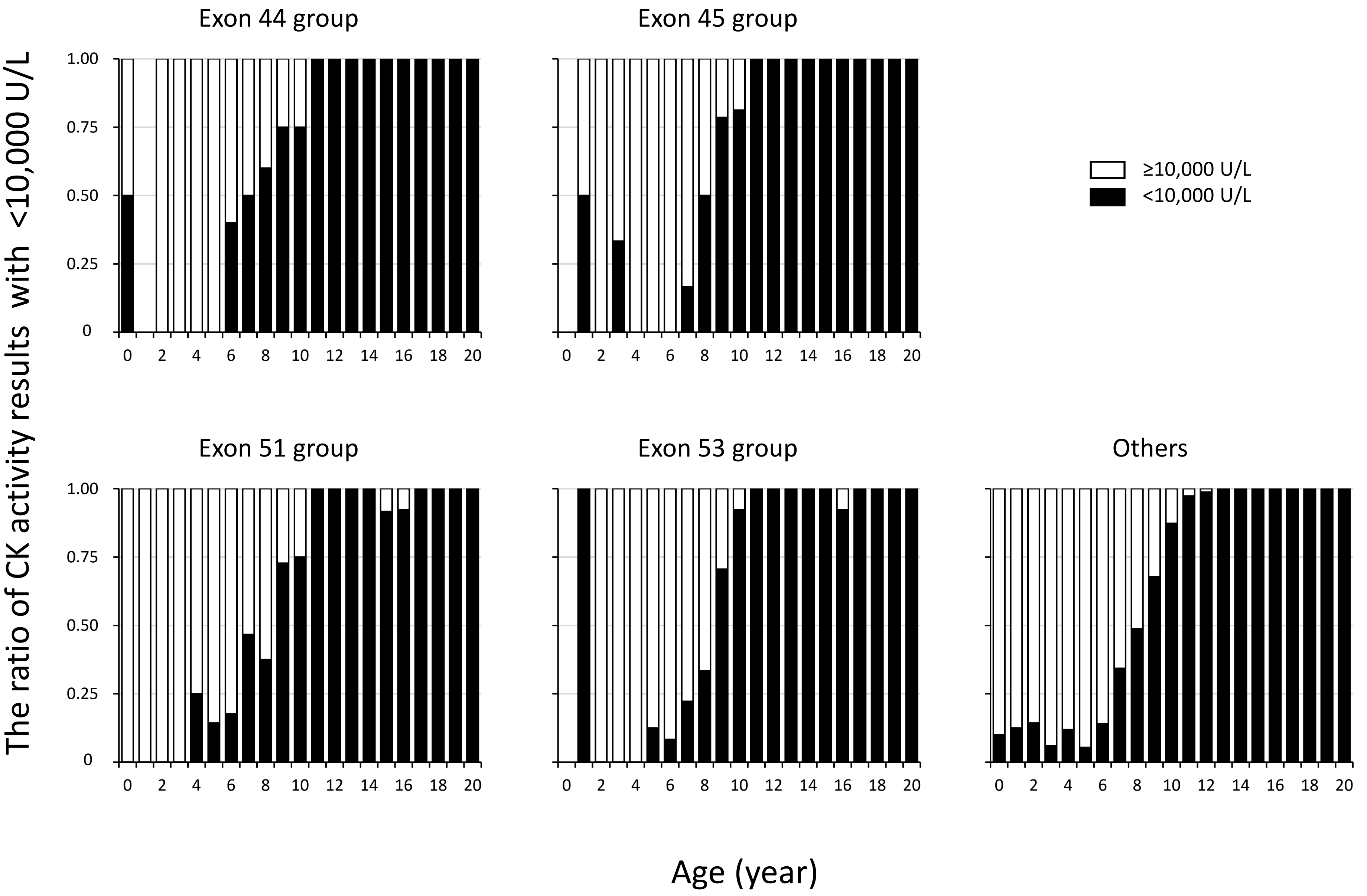
Exon 44 group
Exon 45 group
Exon 51 group
Exon 53 group
Others
≥10,000 U/L
<10,000 U/L
The ratio of CK activity results with <10,000 U/L
1.00
0.75
0.50
0.25
0
0 2 4 6 8 10 12 14 16 18 20
Age (year)

A)

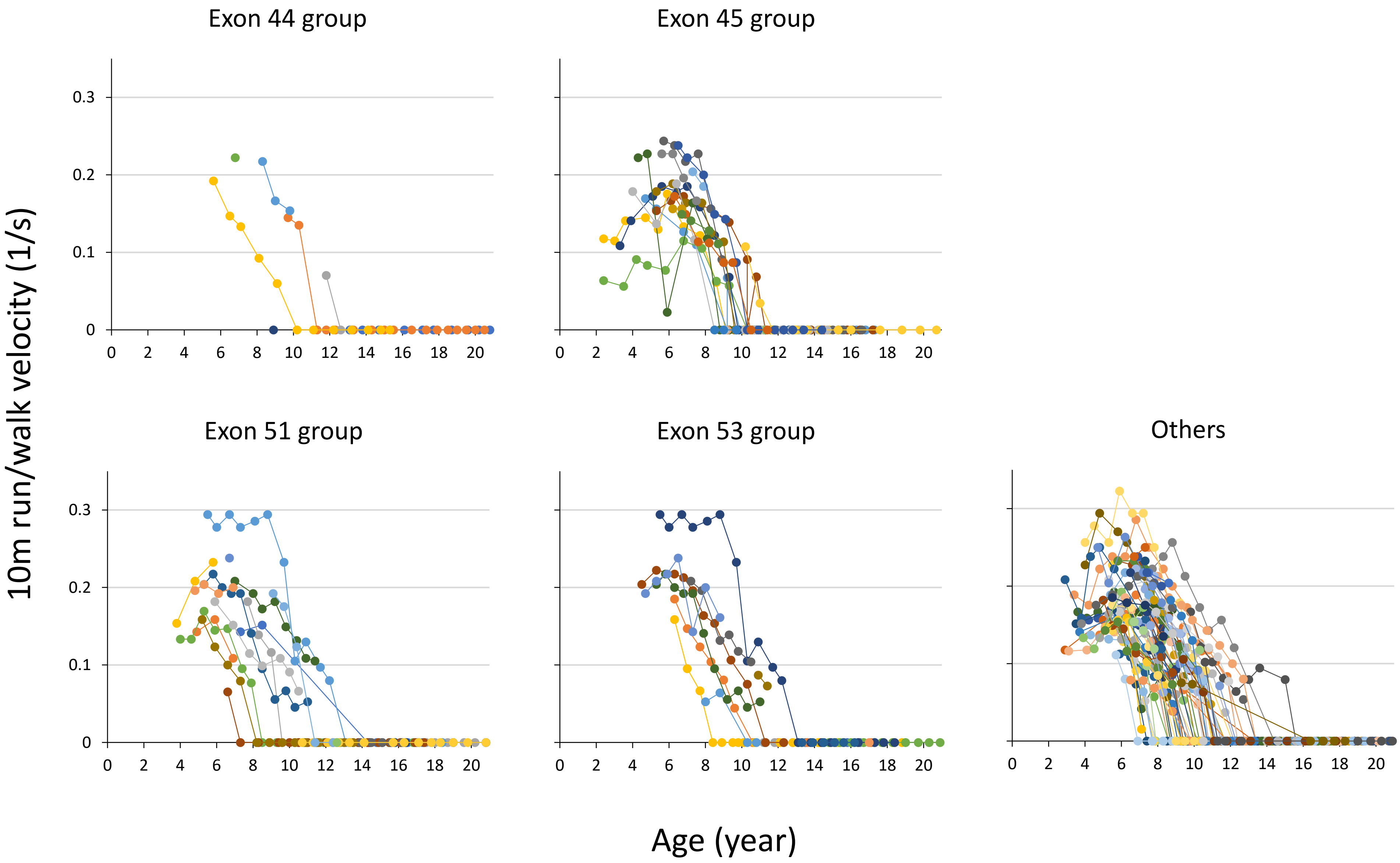

Exon 44 group
Exon 45 group
Exon 51 group
Exon 53 group
Others
10m run/walk velocity (1/s)
Age (year)
0
0.1
0.2
0.3
0
2
4
6
8
10
12
14
16
18
20

B)

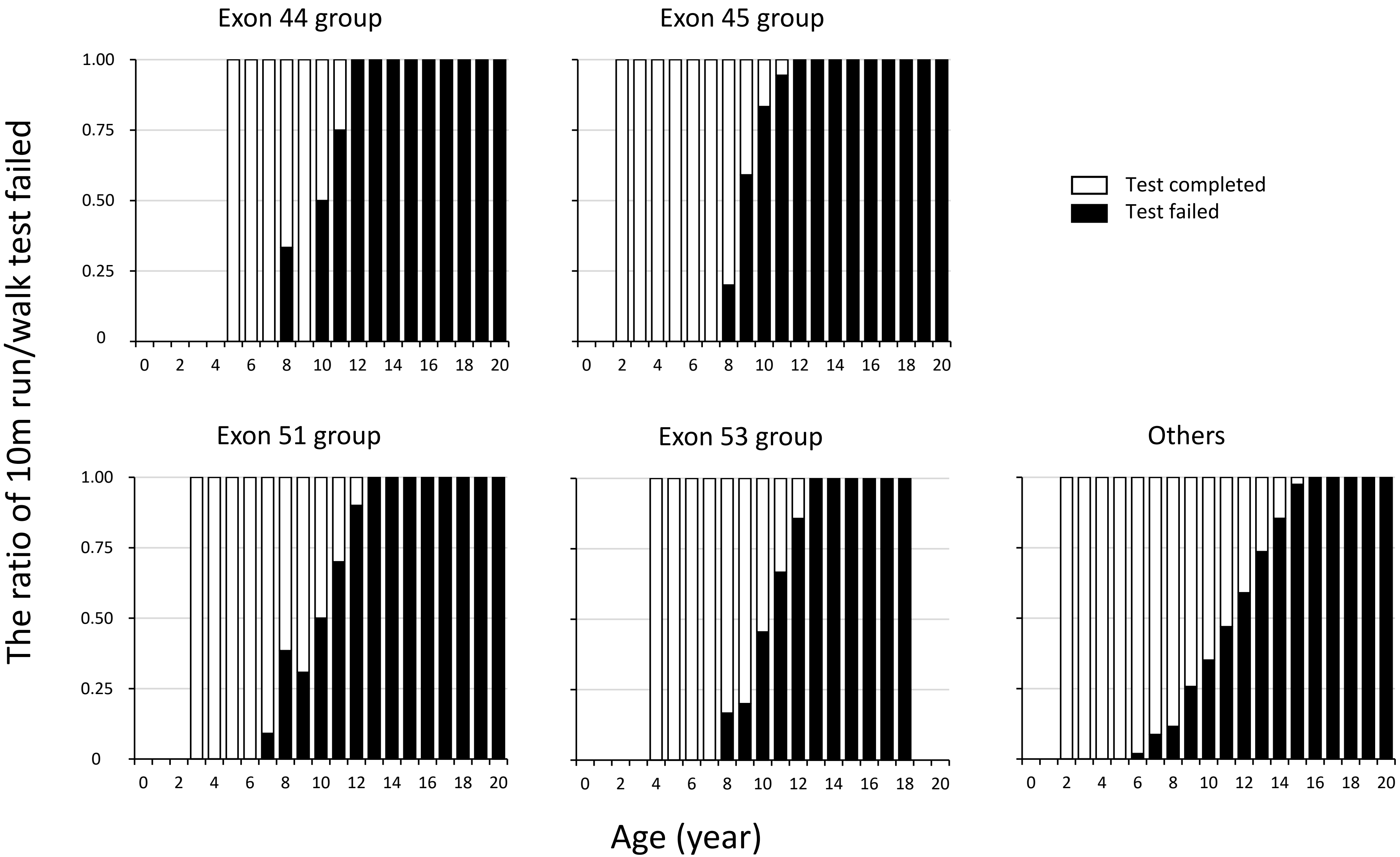

Exon 44 group
Exon 45 group
Exon 51 group
Exon 53 group
Others
Test completed
Test failed
The ratio of 10m run/walk test failed
Age (year)
1.00
0.75
0.50
0.25
0
0 2 4 6 8 10 12 14 16 18 20

c)

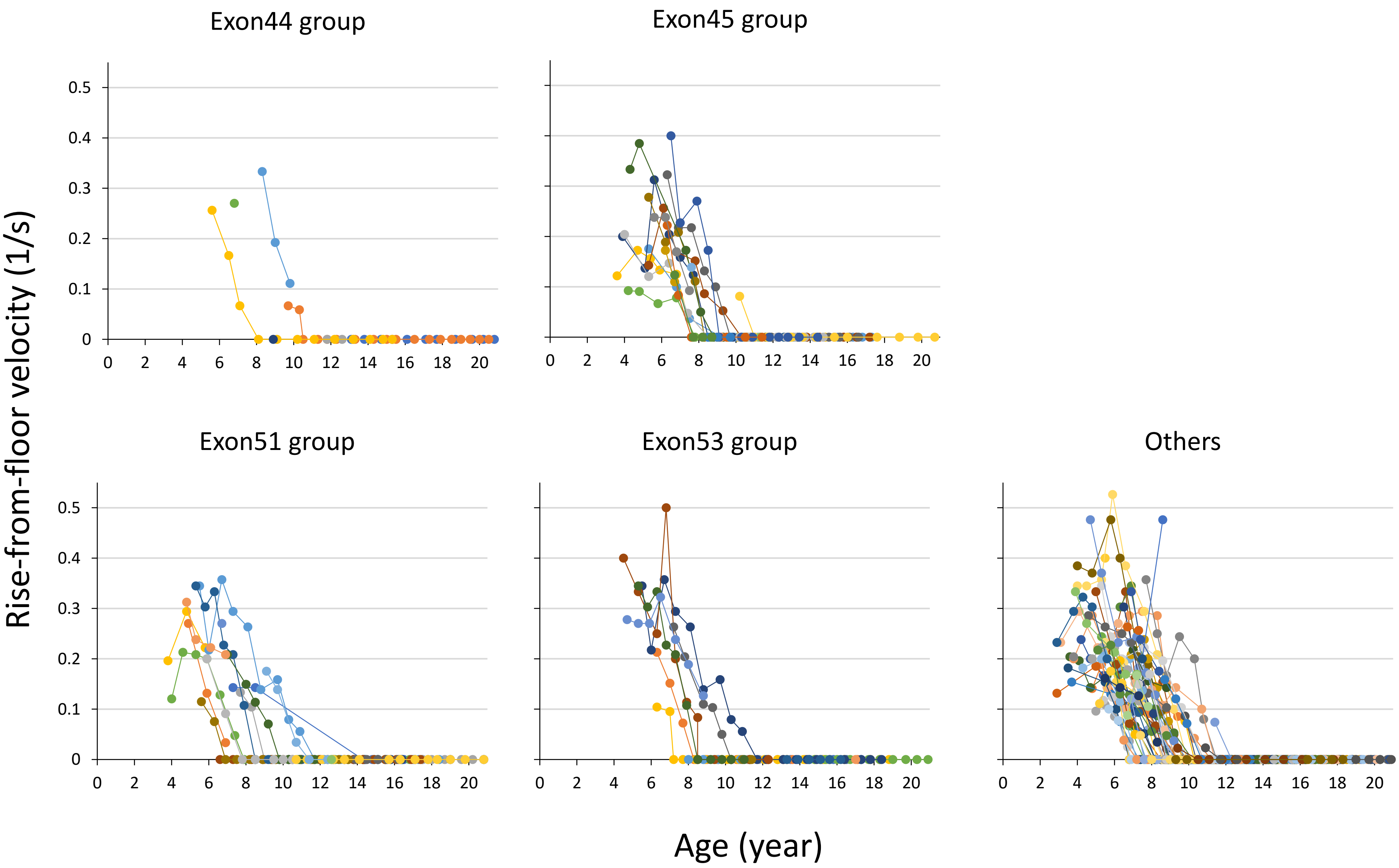
Exon44 group
Exon45 group
Exon51 group
Exon53 group
Others
Rise-from-floor velocity (1/s)
Age (year)
0.5
0.4
0.3
0.2
0.1
0
0 2 4 6 8 10 12 14 16 18 20

D)

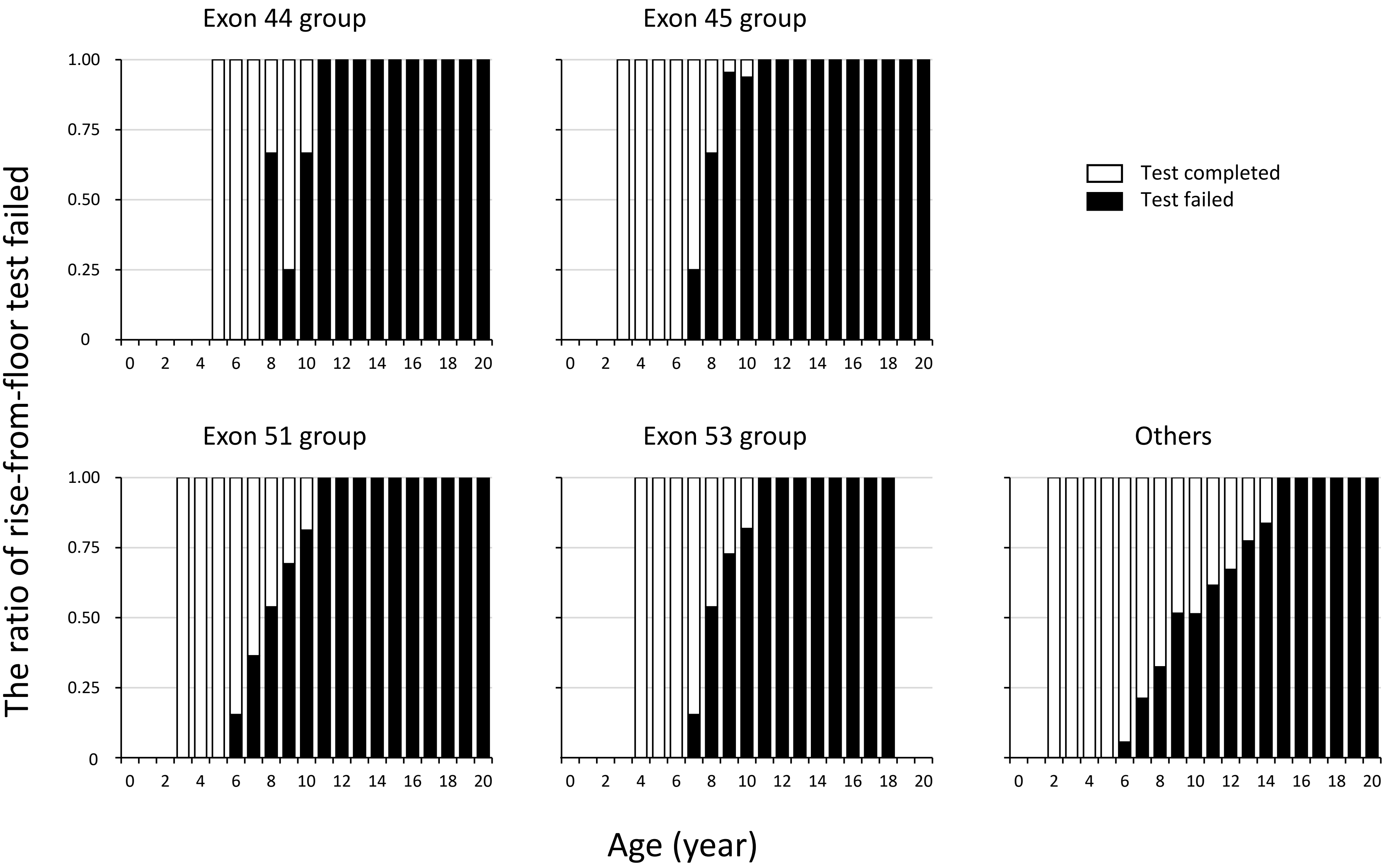
Exon 44 group
Exon 45 group
Exon 51 group
Exon 53 group
Others
Test completed
Test failed
The ratio of rise-from-floor test failed
Age (year)
1.00
0.75
0.50
0.25
0
0 2 4 6 8 10 12 14 16 18 20

A)

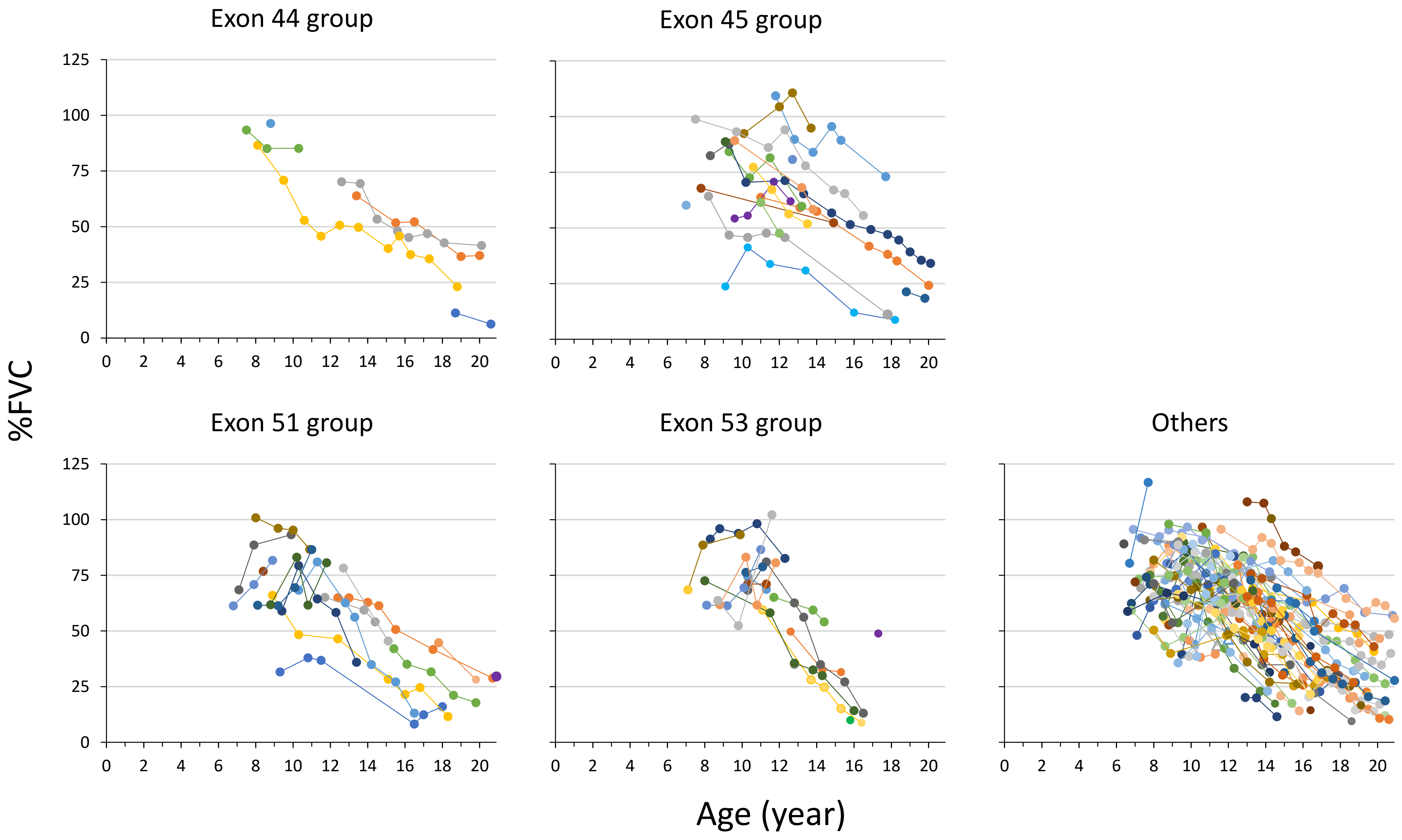

Exon 44 group
Exon 45 group
Exon 51 group
Exon 53 group
Others
%FVC
Age (year)
125
100
75
50
25
0
0 2 4 6 8 10 12 14 16 18 20

B)

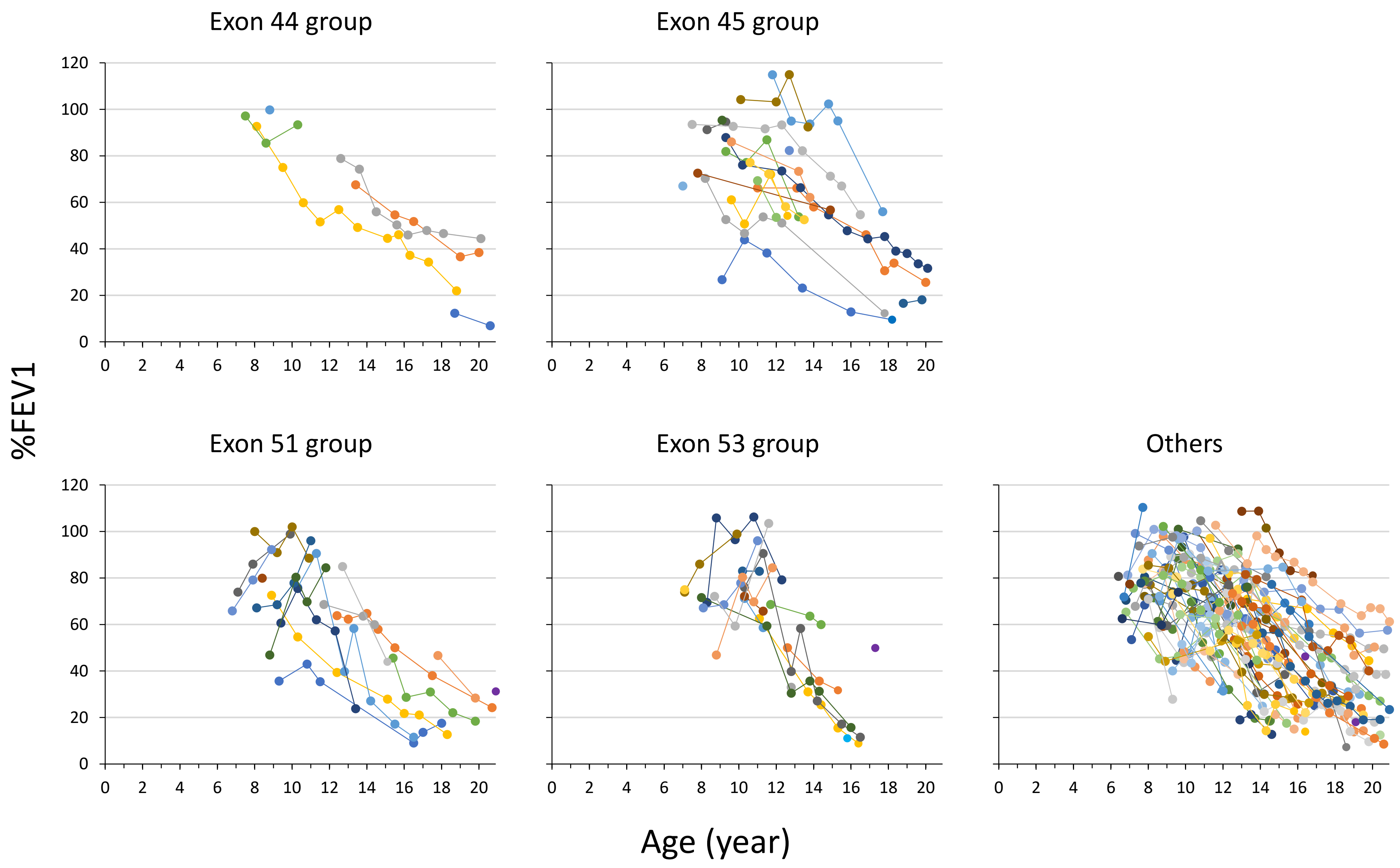

Exon 44 group
Exon 45 group
Exon 51 group
Exon 53 group
Others
%FEV1
Age (year)
120
100
80
60
40
20
0
0 2 4 6 8 10 12 14 16 18 20

C)

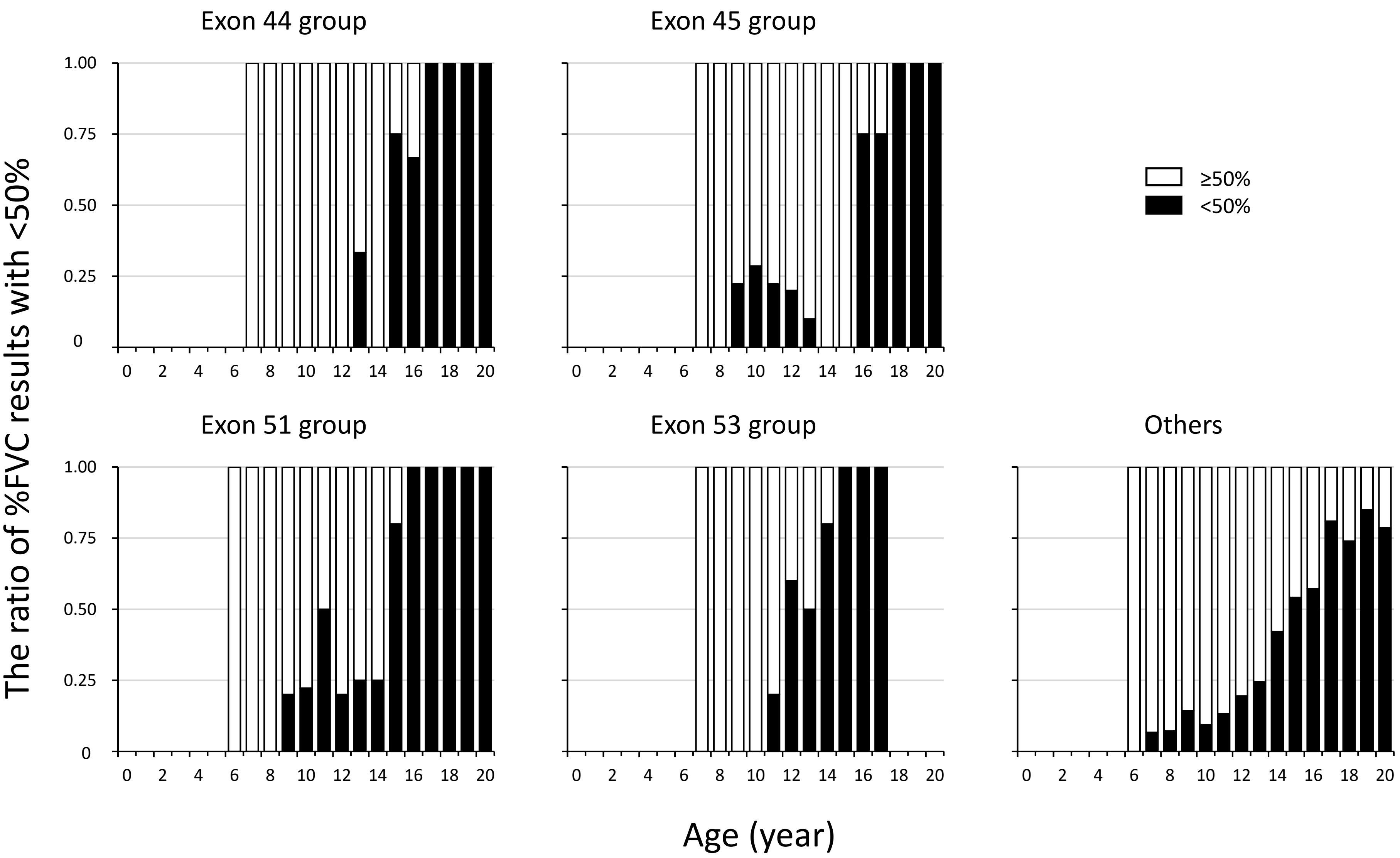

Exon 44 group
Exon 45 group
Exon 51 group
Exon 53 group
Others
≥50%
<50%
The ratio of %FVC results with <50%
Age (year)
1.00
0.75
0.50
0.25
0
0 2 4 6 8 10 12 14 16 18 20

A)

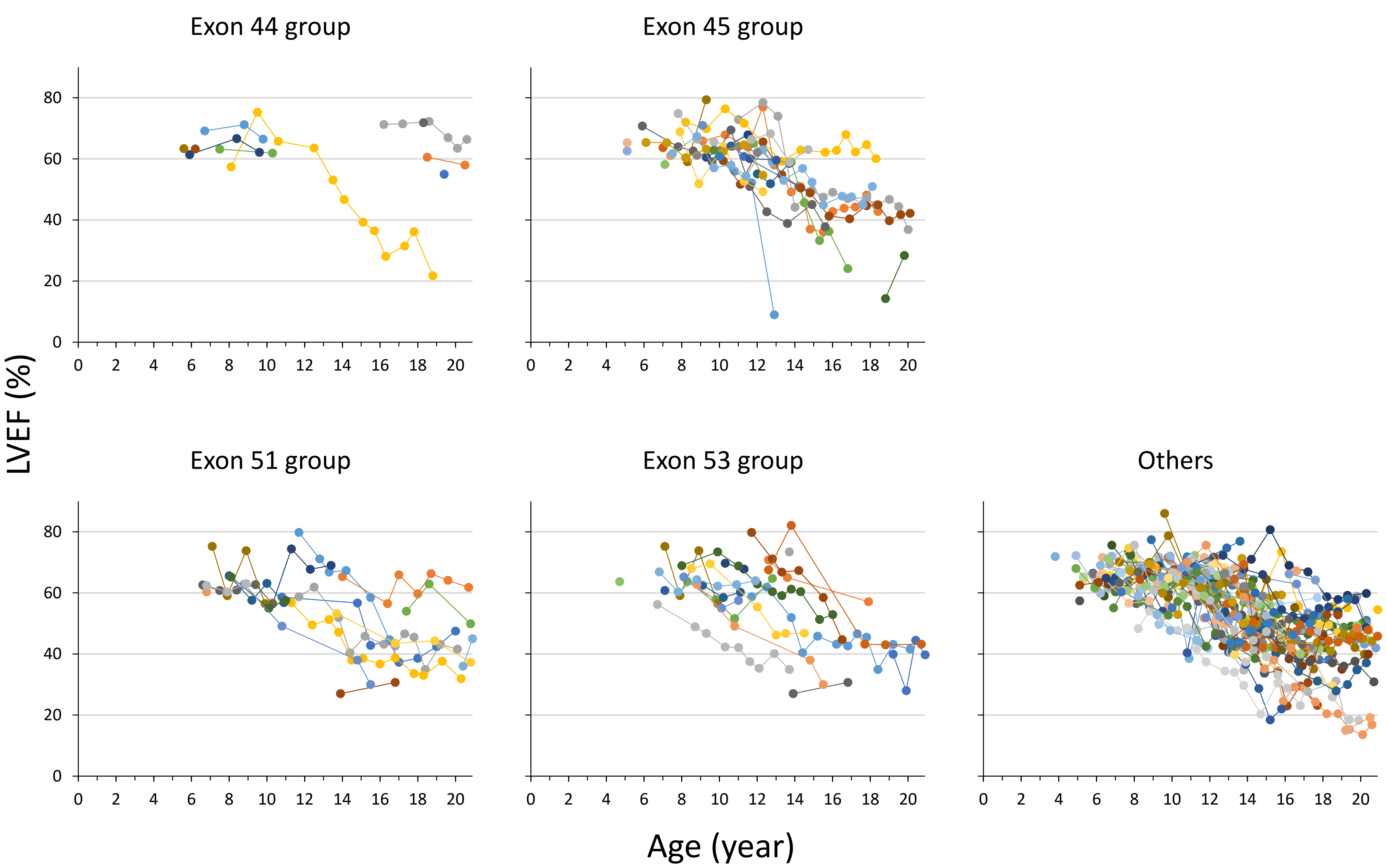

Exon 44 group
Exon 45 group
Exon 51 group
Exon 53 group
Others
LVEF (%)
Age (year)
0
20
40
60
80
0 2 4 6 8 10 12 14 16 18 20

B)

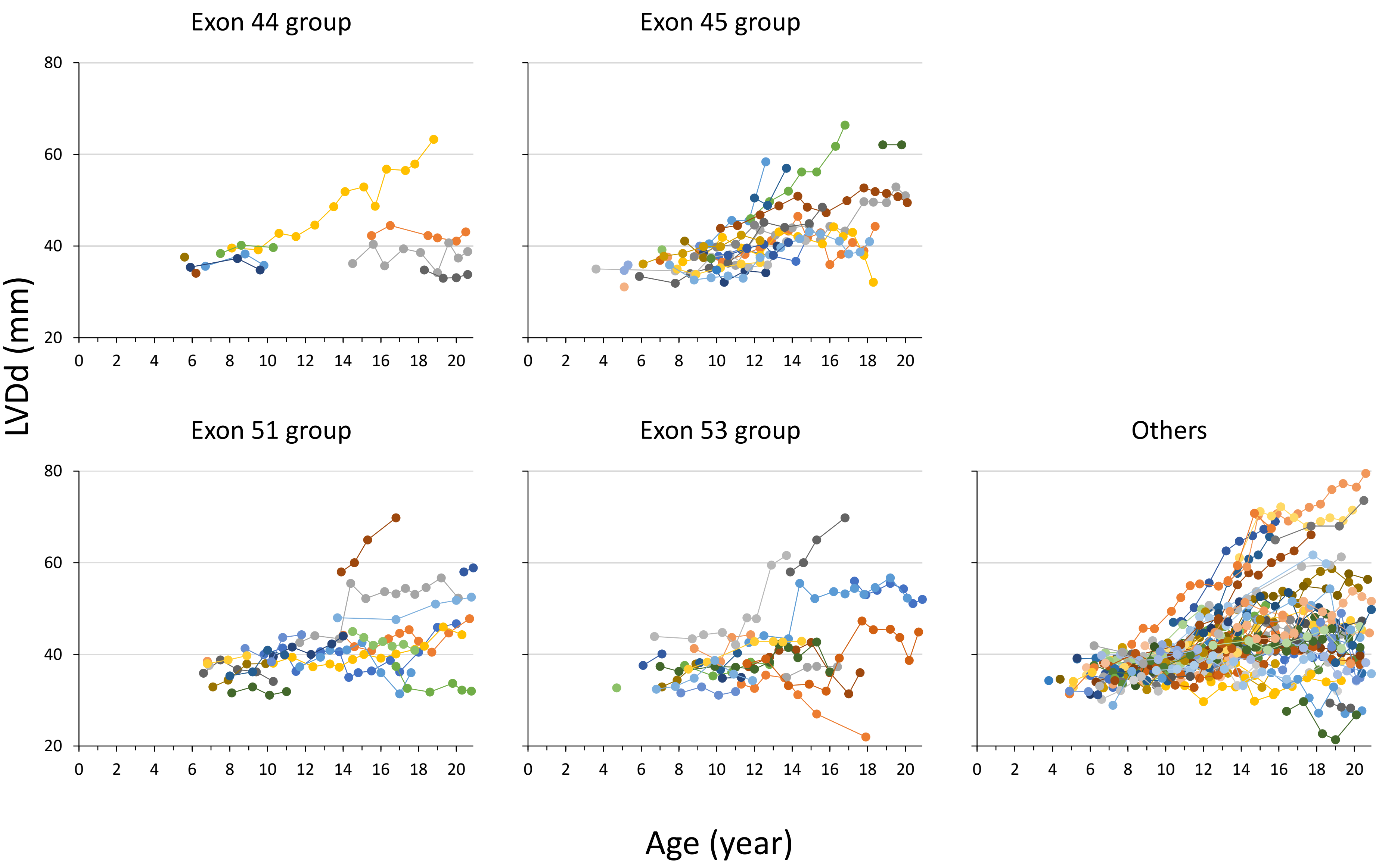
Exon 44 group
Exon 45 group
Exon 51 group
Exon 53 group
Others
LVDd (mm)
Age (year)
80
60
40
20
0
2
4
6
8
10
12
14
16
18
20

c)

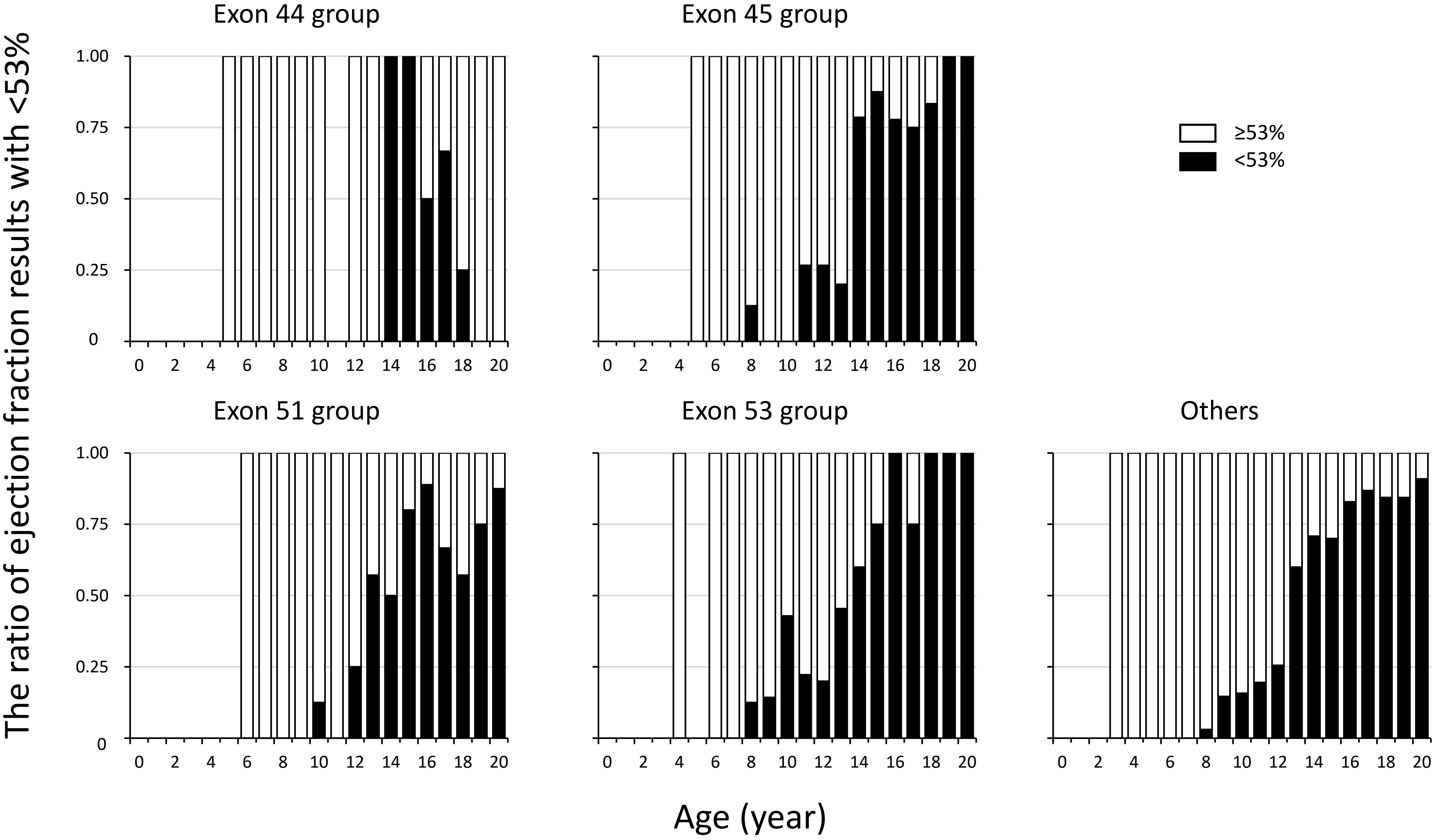

Exon 44 group
Exon 45 group
Exon 51 group
Exon 53 group
Others
≥53%
<53%
The ratio of ejection fraction results with <53%
Age (year)
1.00
0.75
0.50
0.25
0
0 2 4 6 8 10 12 14 16 18 20
